# A TAD-informed aging-brain xQTL atlas of multi-modal and cell-type-resolved regulatory variation

**DOI:** 10.64898/2026.05.21.26353713

**Authors:** Jeffrey Cifello, Ru Feng, Francis P. Grenn, Luke Carter, Anjing Liu, Hao Sun, Ruixi Li, Jenny Anak Empawi, Emily Greenfest-Allen, Zivadin Katanic, Otto Valladares, Amanda B. Kuzma, Heather White, Lindsay A. Farrer, Alison M. Goate, Towfique Raj, Minghui Wang, Carlos Cruchaga, Li-San Wang, Hans Klein, Han Chen, ADSP Functional Genomics Consortium, Philip L. De Jager, Edoardo Marcora, Julia TCW, Xiaoling Zhang, Pavel P. Kuksa, Gao Wang, Yuk Yee Leung

## Abstract

Understanding the regulatory consequences of genetic variation in the aging human brain requires molecular maps that span brain regions, cell types and regulatory modalities. We present the Alzheimer’s Disease Sequencing Project Functional Genomics (FunGen-AD) xQTL Atlas, a harmonized resource of molecular quantitative trait loci from four postmortem brain studies, ROSMAP, MSBB, Knight-ADRC and MiGA. The atlas integrates histone acetylation, DNA methylation, gene expression, splicing and protein abundance QTLs across 14 brain regions, 7 major cell types and 17,566 samples, with standardized association, significance-filtered and fine-mapping outputs. To expand discovery beyond conventional 1-Mb *cis* windows, we include variants within Topologically Associating Domains (TAD) and their boundaries where appropriate, identifying on average 21% more variant-molecular-trait associations per dataset. Statistical fine-mapping reduced broad association sets by 95% into credible sets of candidate regulatory variants. Distributed through the NIAGADS xQTL portal and bulk-download services, the atlas provides a comprehensive functional-genomic foundation for interpreting genetic risk variants in Alzheimer’s disease and aging-brain research.

## Introduction

Most disease-associated variants lie outside of protein-coding sequences and do not directly implicate genes or mechanisms. Instead, these variants are thought to influence regulatory programs, often in a context-specific manner that is not captured by association signals alone^1^. Interpreting genetic associations in the aging human brain therefore requires molecular maps that connect variants to genes, molecular traits, brain regions and cell types.

Molecular quantitative trait locus (xQTL) analyses provide a functional bridge by linking genetic variation to molecular phenotypes such as gene expression, splicing, protein abundance and epigenetic states. Our previous xQTLServe Resource^2^ established this model for human brain molecular QTLs by integrating cortical gene expression, DNA methylation and histone acetylation QTLs with downloadable statistics and browser-based access^2^. It showed that brain xQTL maps can function as community resources for annotating disease-associated variants and prioritizing downstream functional studies.

Since that first-generation brain resource, the data and analytical landscape have changed substantially. Among resources developed or substantially expanded after xQTLServe^2^, the Genotype-Tissue Expression (GTEx) project established a foundational catalogue of expression QTLs across human tissues^3^. Subsequent resources, including the eQTL Catalogue ^4^, MetaBrain^5^, xQTLAtlas^6^ and a recent meta-analysis of single-nucleus eQTL^7^ expanded both the scale and scope of QTL mapping. These studies have incorporated additional molecular traits and increased tissue coverage, providing a growing reference for regulatory variation.

For aging-brain genetics, the remaining need is not simply more QTL associations but resources that combine disease-relevant datasets, multiple molecular modalities, single-nucleus cell-type resolution, consistent allele and effect-size representation, complete downloadable association files and fine-mapped candidate-variant sets. Existing resources remain difficult to integrate and interpret in practice because differences in analytical pipelines, file formats and reporting conventions complicate cross-study comparisons and limit reuse. Many datasets are also distributed as partial outputs or through browser-only interfaces, with inconsistent representation of effect alleles, allele frequencies and effect directions^2–6,8^ (**Supplementary Table S1**).

These limitations are particularly important for Alzheimer’s disease (AD), where genome-wide association studies have identified more than eighty risk loci but the molecular contexts through which most risk variants act remain unresolved^9^. Existing AD-focused QTL studies, spanning bulk tissue, microglia and cerebrospinal fluid, have largely been generated independently, using heterogeneous processing and analysis strategies^8,10–13^. This fragmentation limits interoperability and constrains integrative analyses such as colocalization, Mendelian randomization and fine-mapping.

To address these challenges, investigators in the National Institute on Aging Alzheimer’s Disease Sequencing Project (ADSP) Functional Genomics Consortium (FunGen-AD) established an xQTL Working Group (FunGen-xQTL) to generate a harmonized, brain-centric resource (**Methods**). We present the FunGen-xQTL Atlas, a next-generation human brain xQTL resource spanning histone acetylation, DNA methylation, gene expression, splicing and protein abundance QTLs across fourteen brain regions and seven cell types from four large postmortem brain studies, including the Religious Orders Study and Memory and Aging Project (ROSMAP), Mount Sinai Brain Bank (MSBB), Charles F. and Joanne Knight Alzheimer’s Disease Research Center (Knight-ADRC), and Microglia Genomic Atlas (MiGA).

All datasets were processed using standardized pipelines with stringent quality control and harmonized through a unified framework developed in collaboration with the National Institute on Aging Genetics of Alzheimer’s Disease Data Storage Site (NIAGADS), enabling consistent representation of variant-level associations across cohorts and molecular phenotypes (**Methods**). The standardized protocol is itself part of the resource, providing a reusable infrastructure for future xQTL releases from harmonized genotype and molecular data to association, fine-mapping and dissemination-ready outputs. The same framework incorporates topologically associating domain (TAD)-boundary-enhanced *cis* definitions where appropriate to capture regulatory relationships beyond conventional 1-Mb *cis* windows and applies statistical fine-mapping to prioritize candidate regulatory variants.

Each dataset is released with structured summary statistics and rich metadata, addressing limitations of non-standard outputs common in prior studies^14^. To facilitate access and reuse, all results are available through the project website (https://xqtl.niagads.org), which supports searching, browsing and visualization of xQTLs across genes, variants and genomic regions. The portal incorporates the NIAGADS GenomicsDB genome browser^15^ for interactive exploration of QTLs across molecular phenotypes, tissues, cell types and cohorts. Data are downloadable via the NIAGADS Open Access Data Portal^16^ and the AD Knowledge Portal^17^. Together, these resources support interactive xQTL lookup and downstream statistical integration, including bulk reuse of standardized summary statistics, fine-mapped regulatory-variant prioritization and GWAS-based analyses that require harmonized xQTL effects.

## Results

### The FunGen-xQTL Atlas across cohorts, molecular modalities and cell types

We constructed the FunGen-xQTL Atlas by integrating QTLs derived from multiple molecular phenotypes across four cohorts, ROSMAP^12,18–21^, MSBB^22,23^, Knight-ADRC^8,10,24^, and MiGA^13^ (**Supplementary Methods**). These datasets span fourteen brain regions and seven major brain cell types (**Table 1**) and underwent standardized processing and harmonization utilizing hipFG^25^, enabling consistent representation of genetic variants and QTL associations across studies. The resulting harmonized summary statistics were contributed to NIAGADS^22^ and are publicly available (**Data availability**).

**Table 1.**
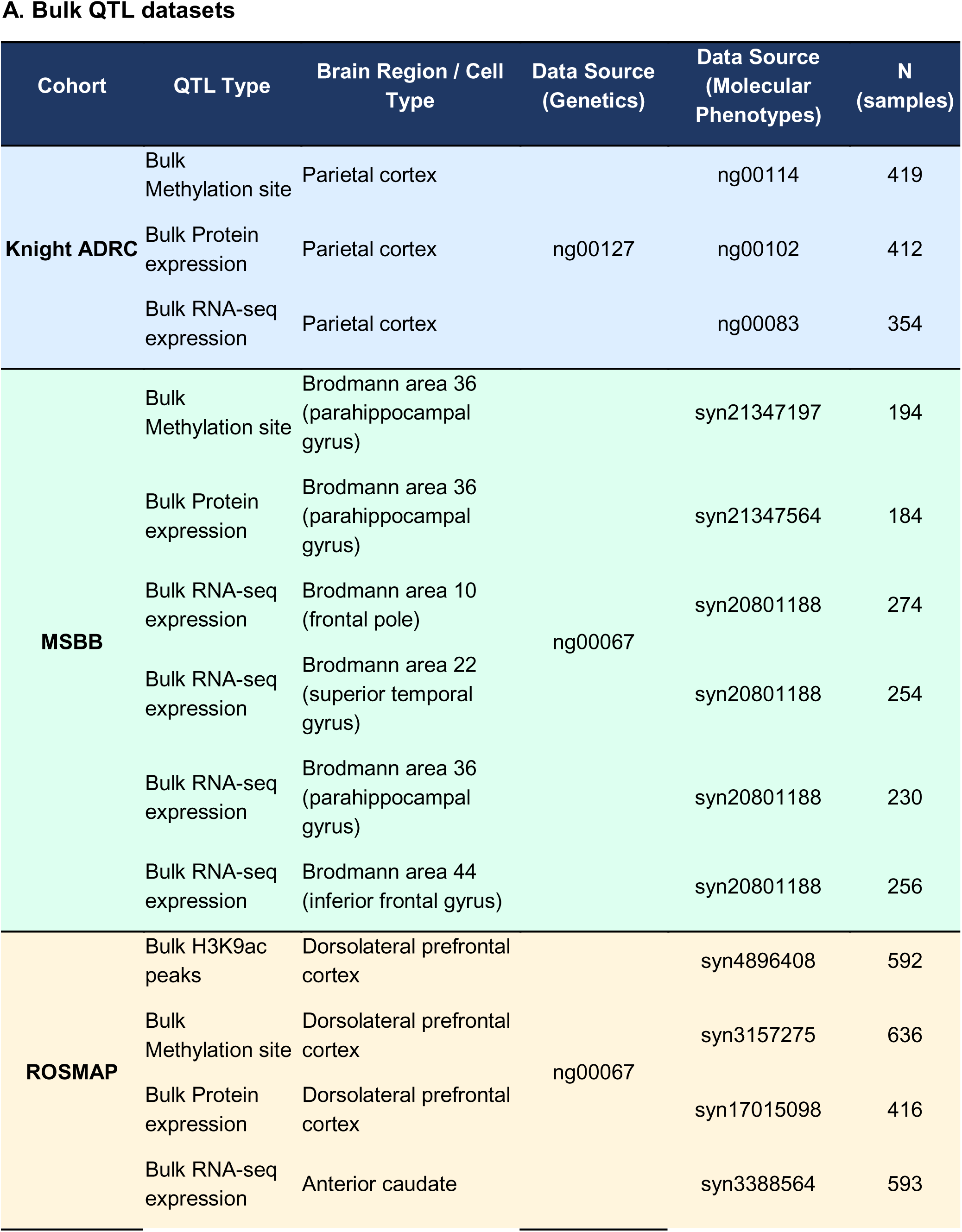
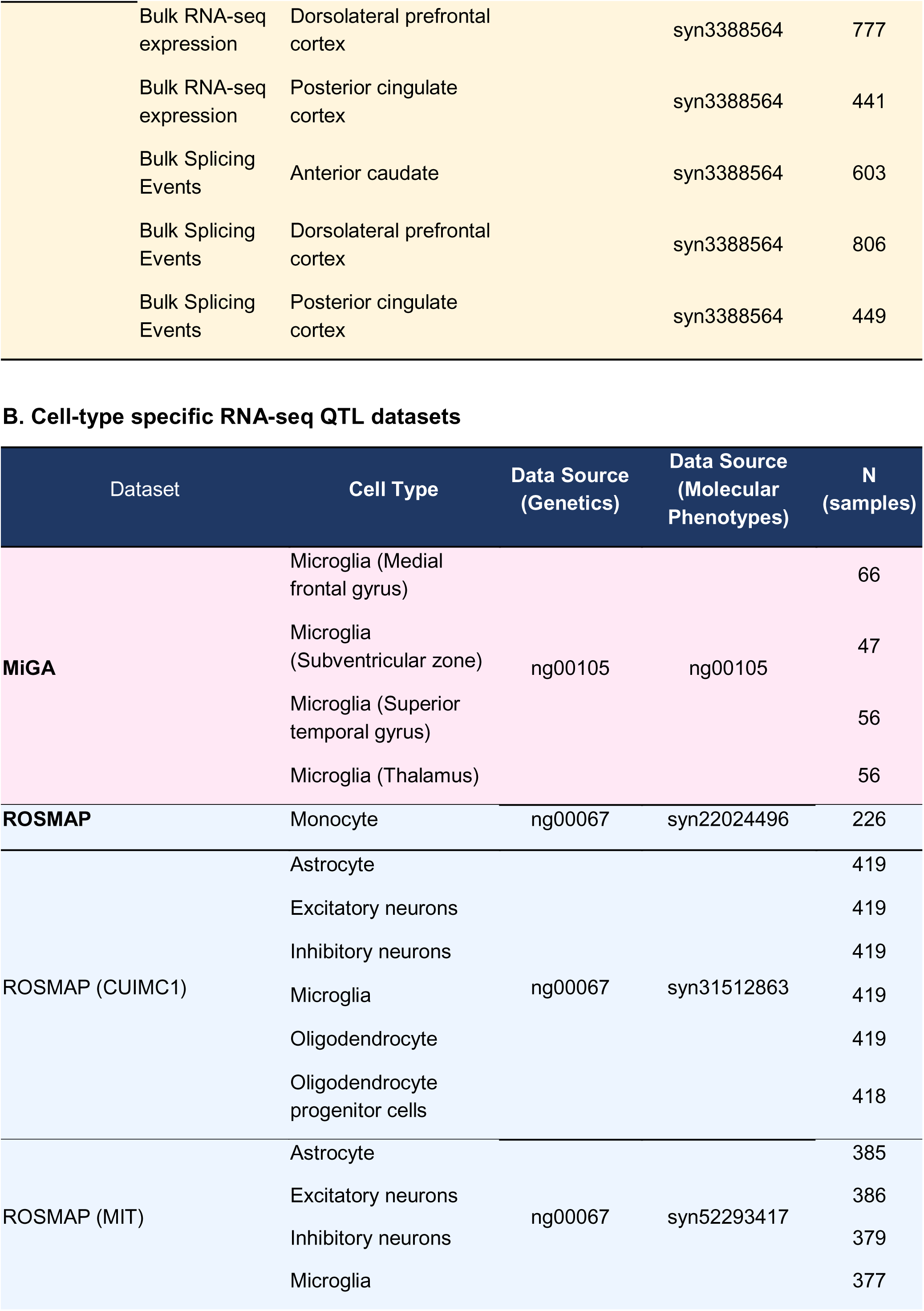
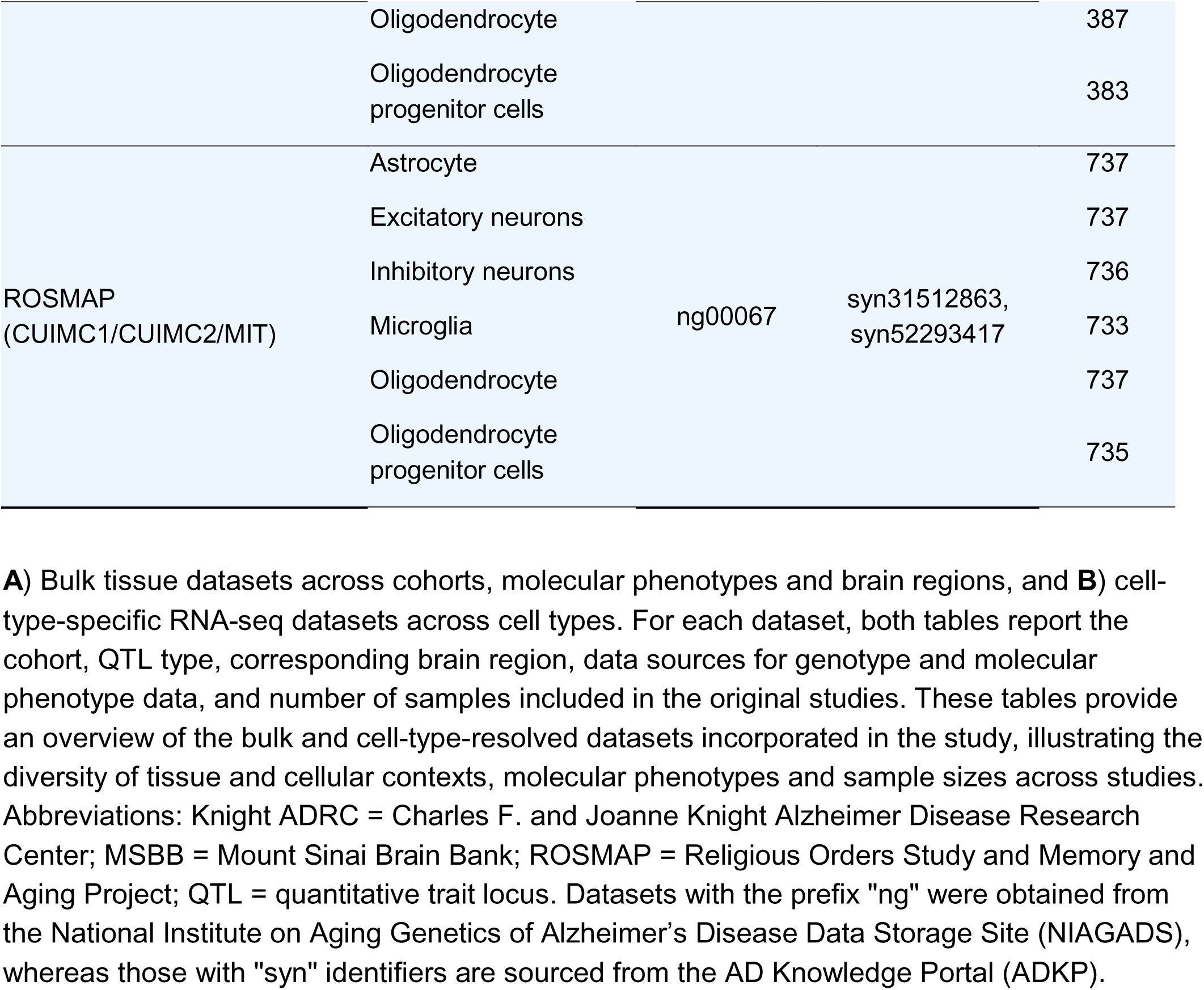
Overview of cohorts, molecular phenotypes, and tissue- and cell-type contexts in the xQTL datasets.

The atlas includes six QTL types, gene expression (bulk eQTL and single-nucleus eQTL, hereafter snuc-eQTL), splicing (sQTL), protein abundance (pQTL), DNA methylation (mQTL), and histone acetylation (haQTL). In total, 17,566 samples are represented across cohorts, capturing both tissue-level (**Table 1A)** and cell-type-resolved regulatory variation (**Table 1B**).

The atlas design and data flow are summarized in **Fig. 1A**, with detailed processing, association-testing and harmonization workflows provided in **Methods**, **Supplementary Fig. S1** and **Supplementary Note 1**. The breadth of molecular phenotypes, brain regions and cell types represented across the atlas is shown in **Fig. 1B** and **Table 1**. The eQTL component builds largely on established GTEx^3^ and TOPMed eQTL^26^ protocols, whereas other QTL layers follow assay-specific best-practice processing, association testing and harmonization steps (**Methods**). Standardized preprocessing and quality control across datasets provide a common framework for comparing xQTL signals across cohorts, molecular phenotypes and biological contexts.

**Figure 1.**
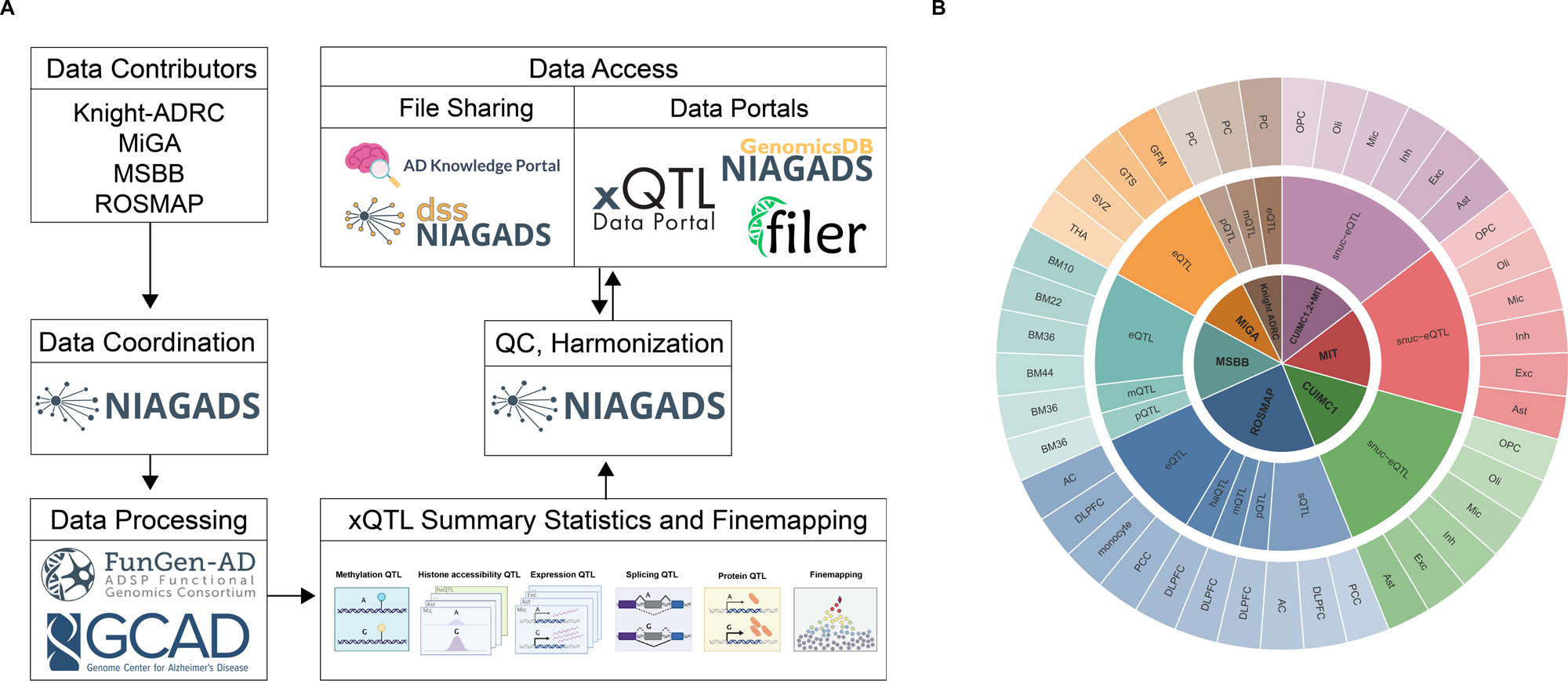
Overview of xQTL data integration and processing. **A)** Overview of the xQTL data generation, harmonization and dissemination pipeline. Multiple studies (Knight-ADRC, MiGA, MSBB and ROSMAP) contributed raw genetics, multi-omics and phenotype data, with xQTL data generation, data sharing and coordination led by NIAGADS (**Methods**). Data were processed using newly developed FunGen-AD and NIAGADS pipelines to generate QTL summary statistics, including methylation (mQTL), histone acetylation (haQTL), expression (eQTL), splicing (sQTL) and protein QTLs (pQTLs), followed by statistical fine-mapping (**Methods** and **Code Availability**). All datasets were processed through the standardized NIAGADS quality-control and harmonization workflow developed for this project (**Methods; Supplementary Methods; Code availability**). Resulting summary statistics and metadata are distributed via file-sharing platforms, including the NIAGADS Data Sharing Service (DSS) open access data portal and the AD Knowledge Portal (**Data availability**). **B)** Cross-cohort, multi-omic landscape of xQTL datasets across brain regions and cell types. The inner ring denotes contributing cohorts (Knight-ADRC, MiGA, MSBB and ROSMAP, including ROSMAP-derived single-nucleus (snuc) datasets) (**Supplementary Methods** and **Table 1**). The middle ring represents individual datasets spanning molecular phenotypes (mQTL, eQTL, sQTL, pQTL, haQTL and snuc-eQTL). The outer ring indicates sampled brain regions (dorsolateral prefrontal cortex (DLPFC), posterior cingulate cortex (PCC), parahippocampal gyrus (PHG), anterior cingulate cortex (AC), and Brodmann BA10/22/36/44 areas) and cell types including astrocytes, excitatory and inhibitory neurons, microglia, oligodendrocytes and oligodendrocyte precursor cells (OPCs) (**Table 1**). This panel highlights the breadth of region- and cell-type-resolved xQTL coverage across studies.

### Cross-resource concordance supports atlas quality

To evaluate the robustness and reproducibility of the atlas, we performed two complementary analyses. First, we assessed consistency of xQTL signals within the FunGen-xQTL Atlas across cohorts, tissues, and molecular phenotypes (**Fig. 2A**). Second, we compared bulk eQTL results against established external resources, including GTEx^3^ **(Fig. 2B**) and MetaBrain^5^ **(Fig. 2C**) (**Methods**).

**Figure 2.**
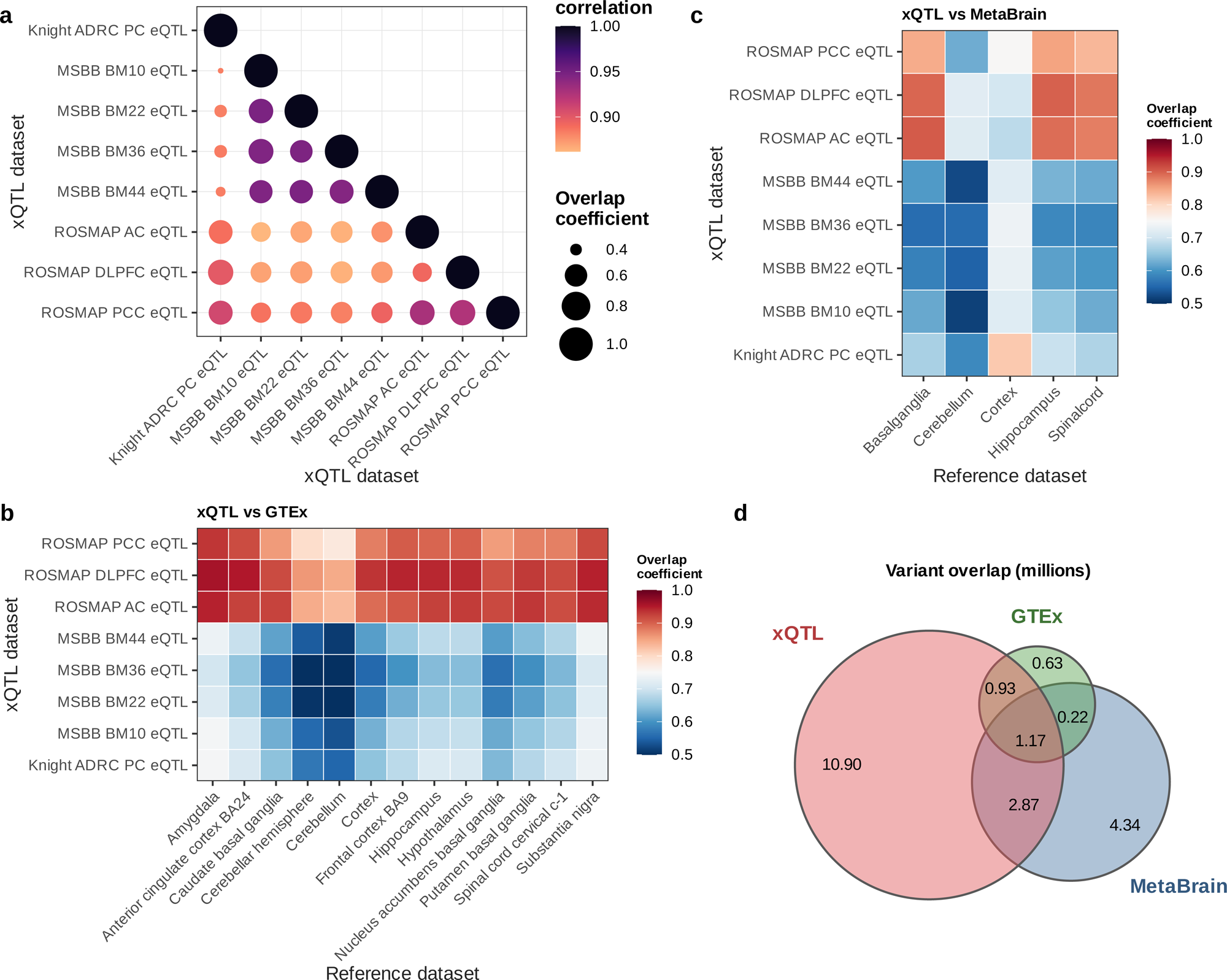
Comparison of the FunGen-xQTL Atlas with existing QTL resources. **A)** Comparison of overlapping associations within FunGen-xQTL Atlas bulk-eQTL datasets, stratified by Spearman correlation coefficient and number of overlapping associations between compared datasets. Spearman correlation coefficients were calculated using z-scores for associations shared between each pair of bulk-eQTL datasets. **B)** Comparison of overlap coefficient (number of overlapping variants between two datasets divided by size of smaller dataset) between FunGen-xQTL bulke-eQTL datasets and GTEx eQTLs (**Methods**). **C)** Comparison with MetaBrain eQTL datasets. We note high overlap (e.g., >0.8 overlap coefficient for ROSMAP eQTL) across all datasets, suggesting that xQTL variant sets cover most of the QTL variants identified in external datasets. **D)** Variant-gene association overlap (Venn diagram) between unique bulk-eQTL variant-gene associations in FunGen-xQTL Atlas bulk-eQTL datasets and other external eQTL resources (GTEx, MetaBrain). Across all contexts, the FunGen-xQTL Atlas datasets capture the majority of unique GTEx brain eQTL associations (71%) and 46% of MetaBrain brain eQTLs, while providing an additional 10.9M novel variant-gene associations not found in GTEx or MetaBrain.

Within the FunGen-xQTL Atlas, we examined reproducibility of detected QTL associations across datasets (except haQTLs, of which there was only one dataset) (**Fig. 2A-C**, **Supplementary Fig. S2**). In the mQTL and sQTL datasets (**Supplementary Fig. S2a, S2b**), Spearman correlations were high for overlapping associations within the same QTL type (0.88-0.98), and we observed strong overlap between the datasets across cohorts and brain regions, with overlap coefficients of 0.4-0.6 for mQTL datasets (**Supplementary Fig. S2b**) and 0.4-0.55 for sQTLs (**Supplementary Fig. S2a**). In the pQTL datasets (**Supplementary Fig. S2c**), we observe comparatively lower Spearman correlations (0.5-0.9) and lower overlap across pQTL datasets from different cohorts, with overlap coefficients of 0.1-0.4; we note that different proteomics platforms were used for ROSMAP and MSBB (**Supplementary Methods**). These results indicate that xQTL signals identified within the atlas are reproducible across independent cohorts, biological contexts, and QTL types, despite the data being generated using different platforms. Overall, replication was highest for brain tissues with similar anatomical context and sample composition, and lower for cross-tissue comparisons, consistent with known tissue specificity of regulatory effects (**Fig. 2A**).

For bulk eQTLs, we benchmarked FunGen-xQTL Atlas results against external resources. We first quantified overlap of significant (FDR<0.05) variant-gene pairs with GTEx^3^ and MetaBrain^5^, observing that 31.2% (4.96M out of 15.9M unique variant-gene pairs) (**Fig. 2D)** of FunGen-xQTL Atlas bulk eQTLs replicated in at least one external dataset.

We also assessed whether the FunGen-xQTL Atlas identifies additional signals beyond those reported in external resources. Many bulk eQTLs (39.8%) (**Fig. 2D)** were unique to the FunGen-xQTL Atlas, reflecting larger and more diverse cohort composition combined with broader coverage of brain regions.

Together, these analyses show that FunGen-xQTL Atlas signals are reproducible both within the atlas and across independent resources. The high degree of concordance, combined with the identification of 2.8-3.3x additional context-specific eQTL associations (33,017,198 bulk-eQTL associations in the FunGen-xQTL Atlas compared to 11,483,598 eQTLs in MetaBrain^5^ and 9,728,785 brain eQTLs in GTEx ^3^), supports the validity of the unified processing framework and the quality of the resulting datasets.

### TAD-boundary-enhanced windows expand distal xQTL discovery

Topologically associating domains (TADs) are genomic intervals with enriched local chromatin contacts^27^, making them a biologically motivated unit for linking distal variants to regulatory targets. We used TAD-boundary-enhanced *cis* regions to extend association testing beyond conventional ±1 Mb windows for expression, splicing and protein traits. Methylation and histone acetylation QTLs used standard ±1 Mb association windows because of computational constraints, but their fine-mapping outputs were harmonized with the other xQTL layers to facilitate downstream integration (**Methods**; **Supplementary Fig. S1**). This design combines generalized brain TADs and boundary regions with gene-centered *cis* windows, producing an average 4.65-Mb analysis window per gene and allowing distal variant-target associations to be tested within regulatory domains (**Fig. 3A**).

**Figure 3.**
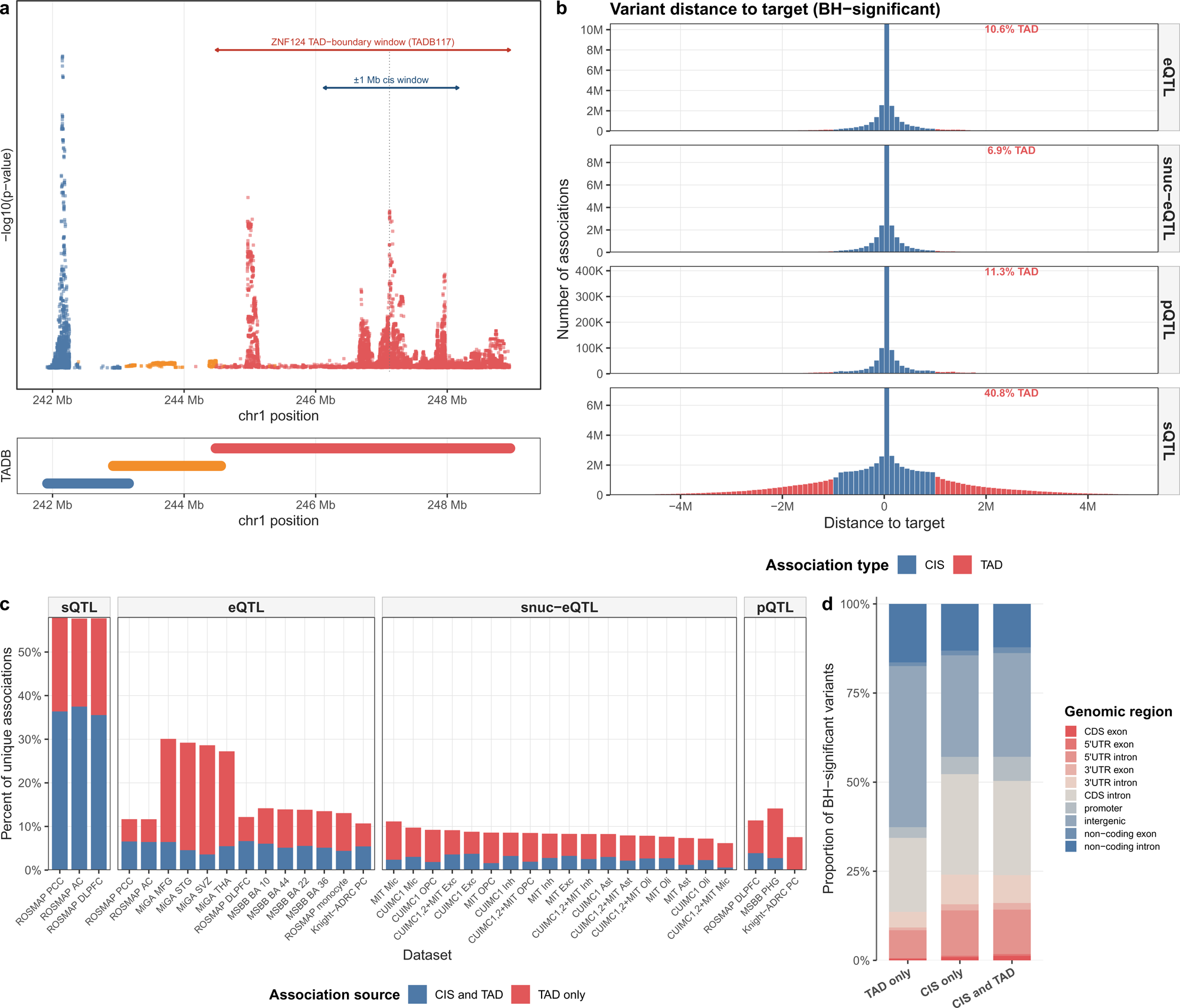
Distribution and genomic context of *cis*- and TAD (>1 Mb) xQTL associations across datasets. **A)** Illustration of TAD-boundary-enhanced *cis* windows (**Methods**). In this example, which utilizes ROSMAP DLPFC eQTL data, we highlight an extended testing window for the ZNF124 gene within TAD-boundary window (TADB117) relative to the traditional ±1 Mb *cis* window, which can yield additional distal associations. **B)** Distribution of genomic distances between associated variants and their molecular targets for Benjamini-Hochberg (BH)-significant associations, stratified by QTL type (**Methods**). Associations are classified as *cis* (red) or TAD-defined distal (>1 Mb) (blue) based on the distance between variant and target. The proportion of TAD (>1 Mb) associations varies by QTL type, with higher fractions of distal associations observed for sQTLs compared to other molecular traits. **C)** Proportion of unique variant–trait associations across datasets and xQTL (sQTL, eQTL, snuc-eQTL and pQTL), stratified by the association distance categories (*cis*-only, TAD-only (>1 Mb), and both *cis* and TAD). Associations are partitioned into variants identified within conventional *cis* windows and those additionally captured through TAD-boundary-enhanced extended windows (>1 Mb). Bars represent the percentage of unique associations per dataset, highlighting variability in the contribution of TAD-resolved signals across cohorts and QTL types. **D)** Genomic location of BH-significant variants (**Supplementary Methods**) across association distance categories from the ROSMAP DLPFC dataset, with all non-snuc-eQTL xQTLs combined. Stacked bar plots show the fraction of variants mapping to functional genomic regions, including promoters, untranslated regions (5′ UTR, 3′ UTR), coding exons and introns, and noncoding exons/introns, demonstrating the predominance of non-coding regulatory regions and the influence of variant-to-target distance on enrichment patterns. cis_and_tad indicates variants with significant associations in both categories; TAD-only and *cis*-only indicate variants found in only one distance category. TAD associations have variant-to-target distances >1 Mb.

Across all datasets, prior to significance filtering, we tested for association 5,584,930,647 bulk eQTLs, 3,390,817,439 snuc-eQTLs, 823,119,430 sQTLs, 309,192,604 pQTLs, 13,982,812,637 mQTLs and 926,340,392 haQTLs, totaling 25,017,213,149 quality-controlled and annotated xQTLs (complete association files (ALL set), **Supplementary Fig. S3, Supplementary Methods**). Following multiple testing correction, 0.3-10.7% of QTLs across QTL types passed Benjamini-Hochberg (BH) false discovery rate (FDR) based filtering (BH-significant files), while 0.1-0.9% were retained under hierarchical multiple testing (HMT) correction (HMT-significant files) (**Supplementary Fig. S3**; **Methods**). These results are summarized in **Supplementary Table S2**.

We then quantified how these expanded windows changed discovery across cohorts and molecular phenotypes (**Fig. 3**). Across datasets, approximately half (48.9%) of all associations originated from variants outside standard 1-Mb *cis* windows and were detected only through TAD-boundary-enhanced *cis* regions (TAD-resolved) (**Supplementary Table S3**). After BH filtering, this proportion decreased to 28.7%, with TAD-resolved (>1 Mb) associations concentrated in sQTLs (40.8% TAD-resolved) and generally lower in the other QTL types (bulk eQTLs: 10.6%, snuc-eQTLs: 6.9%, pQTLs: 11.3%) (**Fig. 3B**). These differences indicate that splicing regulation is more frequently influenced by distal variants located within the same regulatory domain.

The impact of TAD-boundary-enhanced *cis* regions varied across QTL types, by cohorts and by biological contexts (**Fig. 3C**). The three ROSMAP sQTL datasets demonstrated consistently high rates of TAD-resolved associations (41.6-43.7%), but eQTLs demonstrated a wider range (9.7-25.8%), with the MiGA eQTLs all having at least 21.0% of significant associations being TAD-resolved, and the next highest set being 11.5% from ROSMAP AC bulk-eQTLs. pQTLs and snuc-eQTLs had TAD-resolved association rates of 7.4-12.0% and 5.8-10.5%, respectively (**Supplementary Table S3**).

We also assessed the genomic context of QTL variants using ROSMAP DLPFC datasets with all available non-single-nucleus QTL types combined. Most significant variants mapped to non-coding regions, with the strongest non-coding enrichment among TAD-only variants (**Fig. 3D**). Promoter annotations were most frequent among variants detected by both conventional cis and TAD-boundary-enhanced windows, indicating that variants proximal to one target can also contribute to distal regulatory associations. Genome feature distributions were broadly consistent across QTL types and supported the regulatory character of variants identified through TAD-boundary-enhanced regions (**Supplementary Fig. S4**). Together, these analyses show that TAD-boundary-enhanced windows add distal regulatory signals, especially for splicing, while preserving the expected non-coding regulatory profile of xQTL associations. Detailed construction of the TAD-boundary-enhanced analysis is provided in **Methods**.

### Fine-mapping resolves candidate regulatory variants

To resolve candidate regulatory variants for brain molecular QTLs, we applied fine-mapping for protein coding genes across 41 datasets spanning four cohorts and six molecular phenotypes (**Fig. 4A; Methods**) using SuSiE^28^ for gene-related traits (eQTL, snuc-eQTL, pQTL and sQTLs) and fSuSiE^29^ for epigenetic traits (mQTLs and haQTLs). Variant level fine-mapping confidence, as measured by the posterior inclusion probability (PIP), spanned the full range from near-zero to 1.0. **Fig. 4A** shows density of the fine-mapped signals (PIP>0.5) across QTL types and brain tissue/cell type contexts. Fine-mapping substantially reduced the per-target candidate sets of associated variants (BH FDR < 0.05) with 90% of molecular targets across QTL types having a 95% credible set size of only 10% of the BH set (**Supplementary Fig. S5**). Importantly, 24.8% (18,863/76,009) of the fine-mapped variants with PIP > 0.5 across gene expression-related QTL types (eQTLs, snuc-eQTLs, sQTLs, and pQTLs) were TAD (>1 Mb) associations outside of ±1 Mb gene windows.

**Figure 4.**
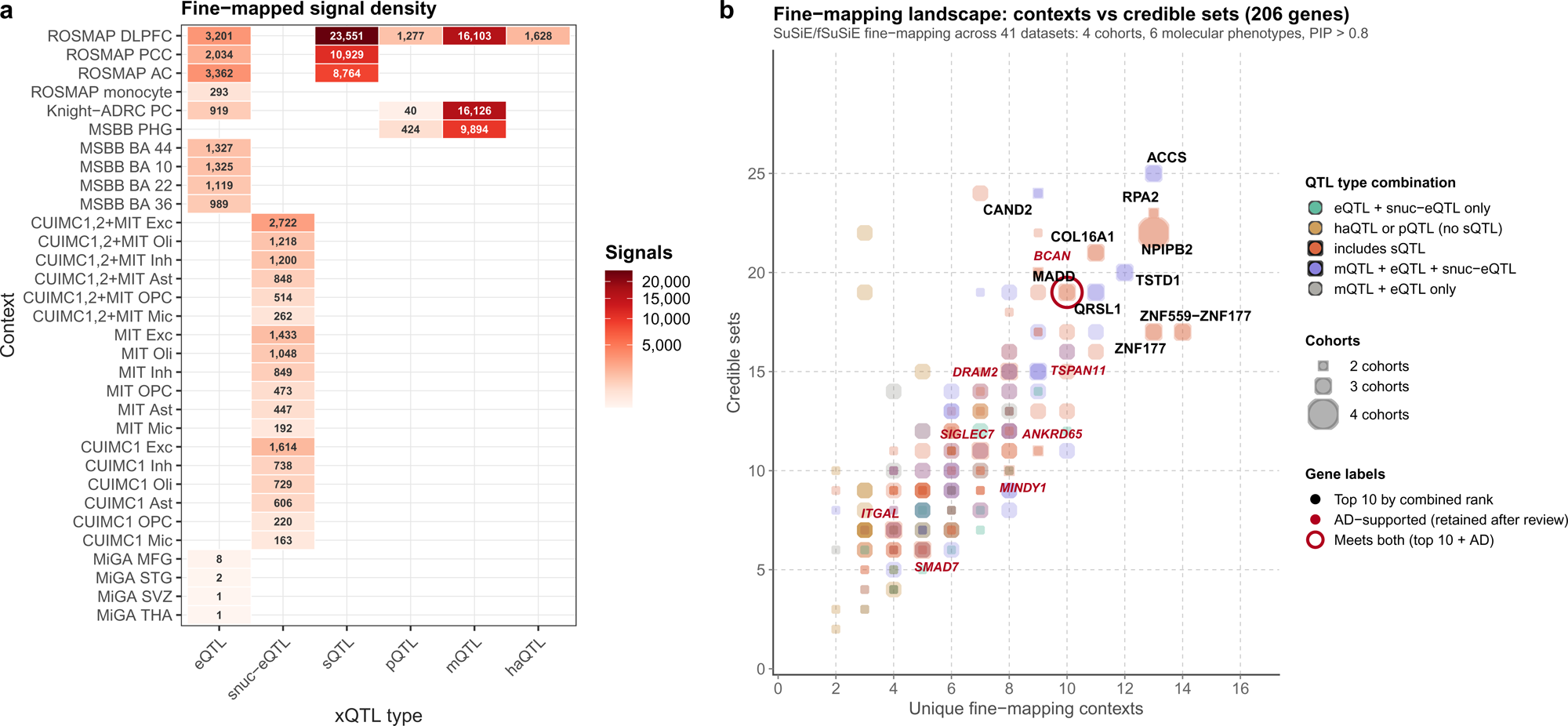
Summary of the fine-mapping data. **A)** Fine-mapped signal density across contexts and xQTL types. The number of 95% credible set fine-mapped variant-molecular phenotype pairs identified for each xQTL type across brain cell types and tissue with PIP > 0.5. Rows represent analysis contexts and columns represent xQTL types, including eQTL, haQTL, mQTL, pQTL, snuc-eQTL, and sQTL. Blank cells indicate context–xQTL combinations that were not available or did not yield fine-mapped signals. The distribution of signals highlights differences in fine-mapped discovery across molecular phenotypes and analysis contexts, with particularly high signal density observed for sQTLs and mQTLs in selected bulk brain regions. **B)** Genome-wide fine-mapping landscape across molecular QTL types and cohorts. Each bubble represents one gene (n = 206) fine-mapped using SuSiE and fSuSiE across 41 datasets spanning four cohorts (ROSMAP, Knight-ADRC, MSBB, MiGA) and six QTL types. The x-axis shows the number of unique fine-mapping contexts, reflecting the breadth of brain region, cell type, and tissue tracks in which a gene harbors at least one 95% credible set with PIP > 0.8. The y-axis shows the total number of credible sets across all active tracks. Bubble color denotes the combination of QTL types with fine-mapping evidence for the gene of interest: orange: at least sQTL; purple: mQTL, eQTL, and snuc-eQTL but not sQTL; teal: eQTL and snuc-eQTL only; amber:haQTL or pQTL but not sQTL; grey: mQTL and eQTL only. Bubble size reflects the number of cohorts contributing fine-mapping evidence. The top 10 genes by combined rank across credible set count and unique context count (summed percentile ranks, lowest = best) are labeled in black: *ACCS, RPA2, CAND2, COL16A1, NPIPB2, TSTD1, QRSL1, ZNF559-ZNF177, ZNF177, MADD* (the last of which also carries AD relevance, see below). Genes with AD-supporting evidence are highlighted in red italics: *BCAN, TSPAN11, DRAM2, ANKRD65, SIGLEC7, MINDY1, ITGAL, and SMAD7*. *ACCS* ranks first on credible sets (n = 25) and *NPIPB2* is the sole gene with fine-mapping evidence across all four cohorts.

To emphasize successfully fine-mapped associations here, we then focused on fine-mapped variants with PIP > 0.5 in gene expression-related traits (eQTLs, snuc-eQTLs, sQTLs, pQTLs, excluding fine-mapping results for epigenetic traits, mQTLs and haQTLs) and required the variant-target pair to be HMT significant in the corresponding xQTL dataset (**Methods**). Following this filter, we identified 95% credible sets for 12,927 protein-coding genes. Across all loci, we identified a total of 59,328 unique fine-mapped variants contained within 74,851 total credible sets. The number of credible sets per gene varied substantially, ranging from 1 (e.g., *TOMM40*,□*KMT2E*) to >20 (*PTPRN2*), reflecting differences in both the complexity of local linkage disequilibrium structure and the breadth of molecular phenotypes affected at each locus.

Out of the 12,927 fine-mapped protein-coding genes, 12,048 harbored at least one high-confidence variant with PIP > 0.8. Of these genes, 1,700 reached PIP > 0.8 in five or more independent QTL tracks, indicative of convergent molecular evidence at those loci. Three genes had PIP > 0.8 variants in 17 or more tracks, *PDPR*, *AK3* and *LRRC37A3*; and 26 genes had such variants in at least 15. Fine-mapping associations for these genes had a range of unique variants (3-29) and credible sets (15-60), demonstrating that highly targeted genes can have varying regulatory networks. For example, *MGMT* had high-confidence PIPs in 29 credible sets but only three unique variants (rs16906252, rs1008982, and rs141539094), with rs16906252 appearing in 27 out of 29, having a minimum PIP of 1.00. Identification of high-confidence signals from across xQTL types and biological contexts allows for gene-prioritization consistent with previous disease-based analyses^30^.

Additionally, 206 genes were found in multiple cohorts, QTL types, and brain contexts with PIP > 0.8 variants in multiple QTL types, cohorts, and brain contexts (**Fig. 4B; Supplementary Table S4**). Nine genes were identified as having known relevance to Alzheimer’s Disease, and one gene, *MADD*, was both a top 10 gene by combined fine-mapping rank and has known AD relevance.

snuc-eQTL tracks, derived from snuc-RNA-seq data across six brain cell types, contributed fine-mapping evidence at 4,083 loci, and were exclusive signals for 644 loci. Notably, *TFG*, *POLE2*, *CCDC59*, *RPN1, CRADD*, *FIGN,* and *SPDYE2* showed high-confidence fine-mapping signals in at least five cell types with no corresponding signals in bulk xQTLs, pointing to cell-type-specific regulatory mechanisms that would be undetectable in bulk tissue analyses.

### Shared xQTLs reveal multi-layer *regulation* at AD risk loci

We quantified the extent to which genetic effects are shared across molecular phenotypes (**Fig. 5**) within the BH-significant datasets. Across the genome, a total of 12,589,636 unique QTL variants were associated with one or more xQTL types and were observed across all autosomes (**Fig. 5A**). To quantify this sharing, we grouped variants by the number of molecular phenotypes they affected. While a large proportion of QTL variants were specific to a single QTL type (28.4%, 3,574,131 variants) (**Fig. 5A**, **top**), a larger fraction of the variants were found to affect two molecular phenotypes (30.74%, 3,870,067 variants), and a combined total of 40.9% (5,145,442) variants were found to affect three or more molecular phenotypes.

**Figure 5.**
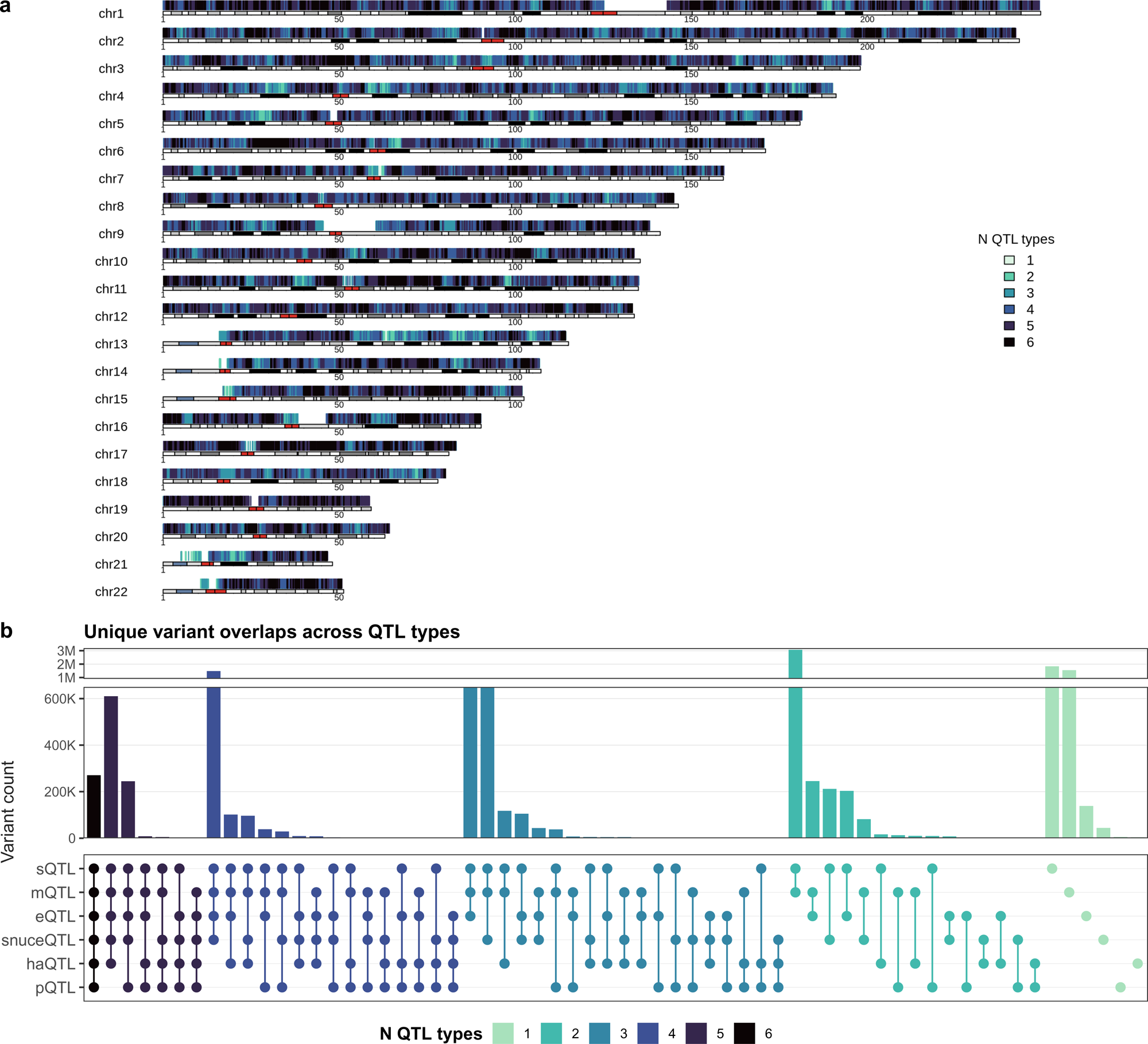
Cross-layer sharing of regulatory variants across molecular phenotype layers. **A)** Genome-wide distribution of xQTL overlaps across molecular phenotypes. Each horizontal track represents a chromosome, with genomic position shown in megabases. Vertical ticks indicate genomic loci harboring variants associated with one or more molecular phenotypes. Colors denote the number of distinct QTL types in which a given variant is detected, ranging from single-type associations to variants associated with multiple QTL types. Variants associated with multiple molecular phenotypes are distributed across all chromosomes, indicating that shared regulatory effects are widespread throughout the genome rather than restricted to specific loci. **B)** Distribution of unique variant overlaps across QTL types, stratified by the number of QTL types in which each variant is involved. **Top**: histogram showing the number of unique variants stratified by the number of molecular phenotypes (QTL types) in which they are detected (from 6 to 1). A substantial proportion exhibit multi-omic effects, with progressively fewer variants shared across higher numbers of molecular traits. **Bottom:** dot-matrix representation of the observed frequencies for combinations of QTL types, including eQTL, sQTL, snuc-eQTL, mQTL, haQTL, and pQTL. Each column represents a unique combination of QTL types, with filled circles indicating inclusion of a QTL type in that combination. This representation highlights both common QTL patterns (for example, eQTL-sQTL combinations) and higher-order combinations spanning multiple regulatory layers.

We next examined the composition of variant overlaps to identify recurring patterns across QTL types (**Fig. 5B, bottom**) within the BH-significant datasets. sQTL-mQTL co-occurrence was the most frequent combination (8,007,923 QTL variants, 63.6% of all 12,589,636 unique QTL variants found across multiple QTL types). While strong overlap existed between bulk and single-nucleus eQTLs (66%, 2,788,124/4,218,357 of snuc-eQTL variants), 34% (1,430,233) of snuc-eQTL variants were found only in single-nucleus datasets and not in bulk eQTL data. Additionally, few variants were distinct to the snuc-eQTL datasets and not found in datasets of other QTL types (1%, 44,228/4,218,357).

As an illustration, we used two reported AD loci^31–33^ to connect these genome-wide sharing patterns to locus-level interpretation, examining BH-significant xQTL signals together with fine-mapping results (**Fig. 6**; **Methods**). At the *ACE* locus, BH-significant associations were observed across eQTL, sQTL, pQTL, mQTL, and single-nucleus eQTL datasets (**Fig. 6A**), spanning multiple tissues and cell types including DLPFC, PCC, AC, and neuronal populations. Fine-mapping separated the *ACE* signals into eleven credible sets containing thirty-three variants across five QTL types and five cohorts. High-resolution splicing and methylation signals mapped to distinct variants, including rs1444621153 (PIP = 1.00) and rs547114353 (PIP = 0.96) for two ROSMAP sQTLs and rs35543459 (PIP = 1.00) for the Knight-ADRC mQTL. Expression-related eQTL, pQTL and single-nucleus eQTL signals were less resolved but directionally concordant, with rs4292 or rs4291 leading the corresponding credible sets.

**Figure 6.**
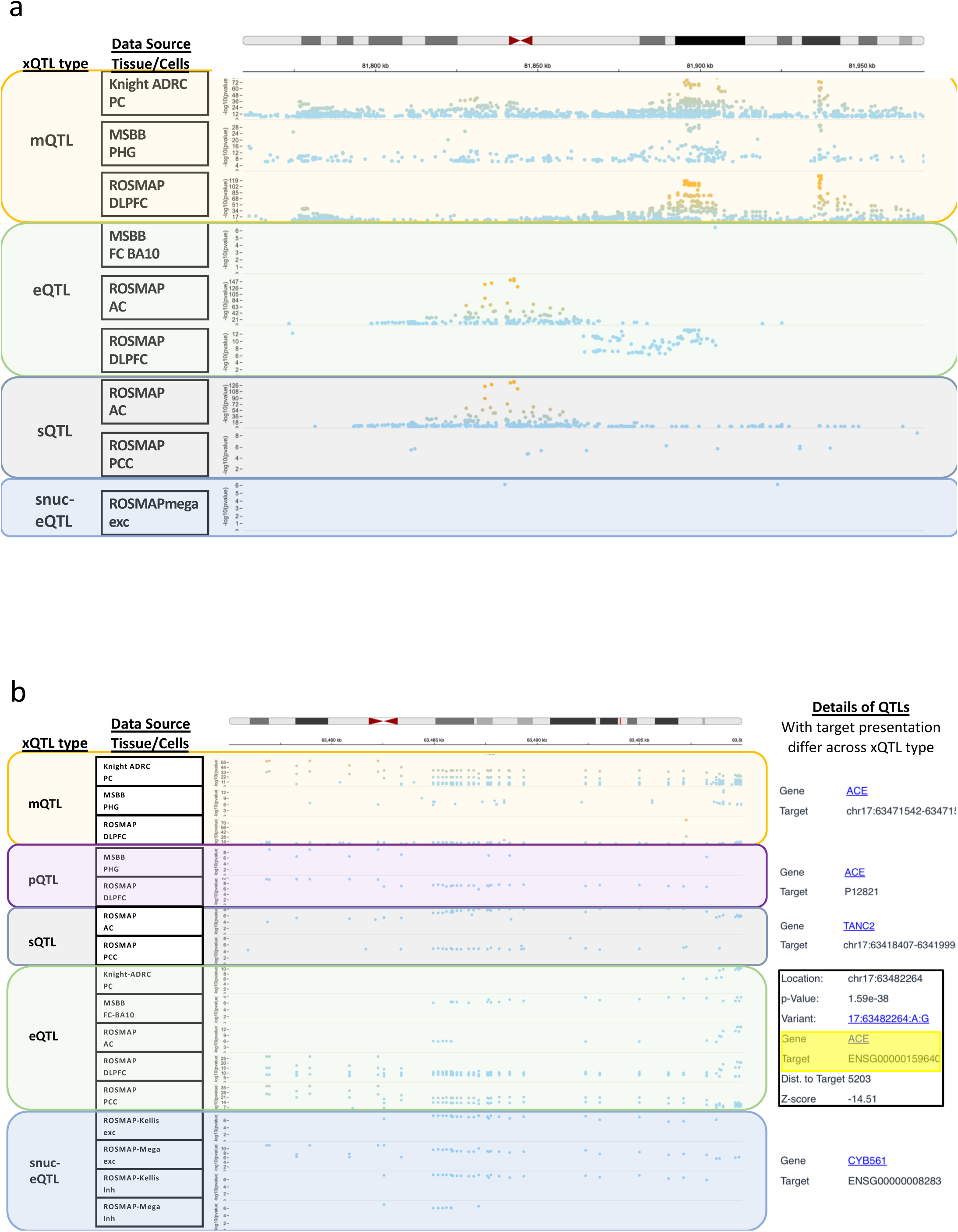
Regional integration of regulatory effects across molecular phenotypes. **A)** *ACE* locus. Regional view of xQTL associations across molecular phenotypes at the *ACE* locus. Tracks display variant–trait associations for mQTL, pQTL, sQTL, eQTL, and snuc-eQTL across cohorts, brain regions and cell types. Each point represents a variant–trait association positioned by genomic coordinate and statistical significance. Associations are observed across multiple QTL types, spanning methylation, protein and transcriptional layers in cortical datasets. Details of xQTL association results are shown for selected entries, including genomic position, p-value, *z*-score, distance to target and target gene assignment; examples of target genes are derived from different variants and reflect QTL-type-specific definitions. **B)** *PLCG2* locus. Regional view of xQTL associations at the *PLCG2* locus across molecular phenotypes. Signals are detected across mQTL, eQTL and sQTL tracks, with distributions varying by dataset and tissue context. Compared to the *ACE* locus, associations are more restricted to specific QTL types, with fewer overlaps across molecular phenotypes. Tracks with fewer than five associations in this region were excluded for visualization purposes. The track-based layout enables comparison of variant–trait associations across datasets within the same genomic interval. Detailed fine-mapping summaries for both loci are provided in **Supplementary Note 2**.

At the *PLCG2* locus, BH-significant associations were observed across mQTL, eQTL, sQTL, and single-nucleus eQTL datasets (**Fig. 6B**), with strong clustering in methylation signals across datasets. Fine-mapping at *PLCG2* identified fourteen credible sets across four cohorts, eleven of which resolved to single variants. Methylation signals were highly resolved, with rs7199820 reaching PIP = 1.00 across three proximal CpG targets in ROSMAP DLPFC, whereas expression signals showed greater allelic complexity across ROSMAP and MSBB contexts. The variant rs8043593 reached PIP = 1.00 in both ROSMAP AC eQTL and sQTL analyses but had opposite effect directions (*z* = +39.7 for eQTL and *z* = -35.8 for sQTL), highlighting distinct regulatory effects on expression level and splicing.

Together, these examples show that loci with similar BH-significant patterns can differ in their underlying regulatory structure. At ACE, signals separate into independent components affecting splicing, methylation and expression-related traits. At PLCG2, many signals resolve to single variants, including one with opposing effects on expression and splicing. Detailed variant-level summaries for these loci are provided in **Supplementary Note 2**.

### Portal and standardized data products enable reuse

The NIAGADS xQTL portal provides a single-entry point for querying, visualization and interpretation of the atlas, together with separately hosted standardized bulk downloads (**Methods** and **Data Availability**). Users can query xQTL associations by variant, gene or genomic region and examine results across molecular phenotypes, tissues, cell types and cohorts (**Fig. 7**).

**Figure 7.**
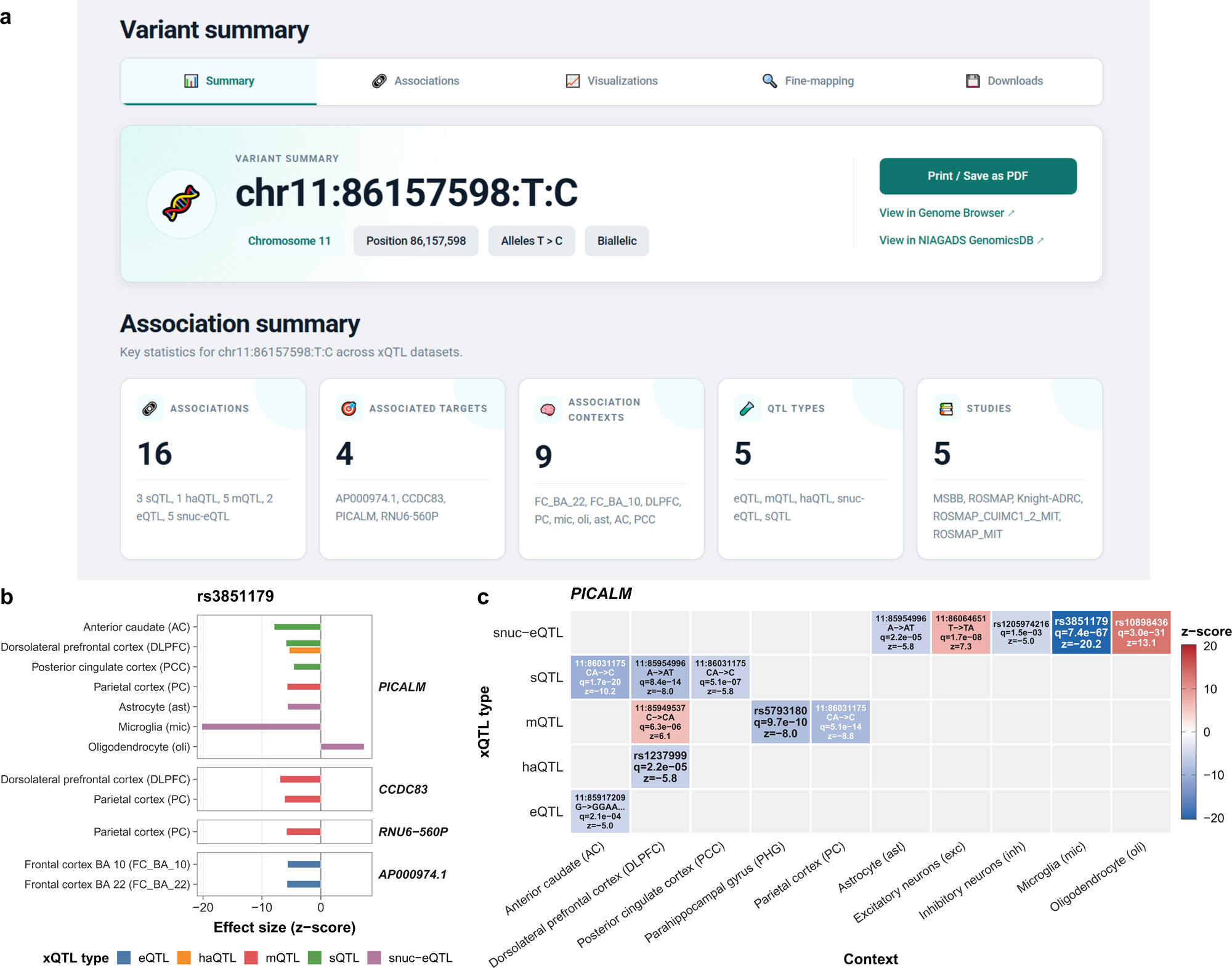
Interactive exploration of xQTL associations through the FunGen-xQTL portal. **A)** Variant-centric summary view for rs3851179, displaying genomic position, allelic information, and aggregated association counts across QTL types, target genes, tissues/cell types, and contributing studies in the FunGen-xQTL portal (https://xqtl.niagads.org; Methods). Summary panels provide direct links to underlying association records and downstream visualizations. **B)** Gene-level and context-resolved effect size view, showing standardized effect sizes (z-scores) for associated targets across brain regions and cell types. Associations are stratified by xQTL type (eQTL, sQTL, pQTL, mQTL, haQTL, and snuc-eQTL), enabling comparison of regulatory effects across molecular layers and biological contexts. **C)** Context-by-QTL type heatmap summarizing the strongest association per cell type or tissue for the selected locus (e.*g., PI*CALM). Color scale represents standardized effect si*z*e (z-score), with annotations indicating variant identity, allele change, false discovery rate (FDR), and direction of effect. This view highlights context-specific and shared regulatory signals across molecular phenotypes.

The interface is organized around a few complementary views. A variant-centric summary (**Fig. 7A**) provides genomic position, allelic information, and counts of associated signals across xQTL types, targets, and studies, with links to underlying records. A context and target view (**Fig. 7B**) displays effect sizes across tissues and cell types for selected genes, enabling comparison of direction and magnitude of associations. A locus-level view (**Fig. 7C**) summarizes association patterns within a genomic region, highlighting the strongest signals across molecular phenotypes and contexts.

In addition to these summaries, users can examine xQTL signals in genomic context through an interactive genome browser powered by the NIAGADS GenomicsDB infrastructure^15^ (**Methods**). This browser supports simultaneous visualization of multiple xQTL tracks and filtering by phenotype, tissue, and cohort. Examples of locus-level views generated through this interface are shown in **Fig. 7**.

All results are released as standardized file sets, including complete association results (ALL), BH-significant associations, HMT-filtered associations and fine-mapping outputs provided as full results and 95% credible sets (**Supplementary Fig. S3**). These files retain harmonized variant representation, QTL statistics and fine-mapping annotations where applicable. The file-set structure and counts across all forty-one datasets are summarized in **Supplementary Tables S2 and S3**, and bulk files are available through the NIAGADS Data Sharing Service^16^ and the AD Knowledge Portal ^17^(**Data availability**).

## Discussion

We present the FunGen-xQTL Atlas as a brain xQTL resource for AD functional genomics. The original xQTLServe^2^ helped establish this resource model by making brain expression, DNA methylation and histone acetylation QTLs searchable, downloadable and usable for genetic-variant interpretation. The FunGen-xQTL Atlas carries this model into current aging-brain genomics by adding splicing and protein QTLs, single-nucleus cell-type contexts, multiple AD-relevant cohorts and standardized association, fine-mapping and portal-ready outputs. Its central contribution is therefore not only a larger catalogue of QTL associations, but a harmonized release in which regulatory effects can be compared across molecular layers, cohorts and biological contexts and reused through NIAGADS^16^.

The atlas is also a framework for producing and distributing comparable functional-genomic resources. Consistent, reusable protocols standardize genotype processing, molecular phenotype quality control, association testing, target annotation and fine-mapping across contributing datasets. The hipFG harmonization framework then converts association and fine-mapping results into a shared summary-statistics format, reducing *post hoc* reformatting and allowing new datasets to be added without changing the data model. The NIAGADS xQTL portal supports variant-, gene- and region-based exploration, while standardized bulk files allow programmatic reuse in downstream analyses. The same protocol and software layer can support future FunGen-AD releases and community groups building comparable xQTL resources, with the portal serving as a durable entry point that can be updated as new datasets are processed. The standardized xQTL outputs can support summary-statistic analyses that combine molecular QTL effects with external GWAS results, including TWAS, colocalization, heritability estimation and related summary-statistic workflows. For methods that require LD information, users should pair the xQTL summary statistics with the companion European-ancestry ADSP/GCAD LD reference panel^34^ (ADSP WGS Release 4, N ≈17,000) available through NIAGADS^16,35^. The same atlas-generation framework provides the upstream molecular datasets used to train the companion molecular phenotype prediction models^36^, linking this upstream xQTL release to downstream TWAS and joint variant-gene fine-mapping resources.

Several analyses in this study support the reliability and added scope of the atlas without requiring users to treat every association equally. Concordance across comparable datasets and with GTEx and MetaBrain indicates that the harmonization framework preserves well-established regulatory signals, while FunGen-xQTL Atlas-specific associations reflect additional brain regions, cohorts, cell types and assays. The TAD-boundary-enhanced design and fine-mapping layer move the resource beyond association discovery by capturing distal regulatory signals within chromatin domains and narrowing broad association sets to candidate regulatory variants with convergent support across contexts. This separation between discovery-scale association catalogs and smaller prioritized candidate sets supports both broad downstream integration using xQTL summary statistics and functional annotation of signals most suitable to guide follow-up functional studies.

The genome-wide sharing analysis and AD risk-locus examples show how the atlas can be used to interpret regulatory mechanisms rather than only to list associations. Across the genome, the atlas distinguishes variants with effects shared across molecular phenotypes from those acting in a specific QTL type, brain region or cellular context. At *ACE* and *PLCG2*, integrated association and fine-mapping views reveal different regulatory architectures, with *ACE* showing broader cross-layer regulation and *PLCG2* showing more selective and allelically complex signals. These examples illustrate why loci with similar genetic association strength can point to different molecular layers and contexts, and the same approach can be applied to additional loci, traits and molecular contexts through the portal or bulk files.

Several limitations should guide use of the FunGen-xQTL Atlas and future releases. Postmortem tissue and cross-cohort heterogeneity introduce variation in sample quality, disease state, assay design and power that cannot be fully removed by covariate adjustment. Cell-type and cell-state resolution remains limited by available sample size, and TAD-boundary-enhanced windows depend on reference chromatin maps that may miss disease-, cell-state- or ancestry-specific regulatory interactions. Fine-mapping narrows candidate variants but remains a statistical prioritization step rather than a causal inference, and downstream use in non-European or mixed-ancestry settings will depend on LD, allele frequency and regulatory-architecture differences. This release should therefore be viewed as a first version of a living FunGen-xQTL Atlas framework rather than a static catalogue. As FunGen-AD continue to generate multi-ancestry data and larger single-nucleus datasets, the same protocol and NIAGADS portal structure can support updates to finer cell subtypes while preserving the multi-modal design of the current resource across expression, splicing, protein abundance, DNA methylation and histone acetylation QTLs. By keeping the data model, quality-control workflow and release structure stable, future versions can remain comparable to the present atlas while expanding its biological and population coverage.

## Methods

### Definitions and resource scope

Throughout the **Methods**, we use molecular dataset to mean a source-specific molecular measurement panel used for xQTL mapping, such as a DLPFC eQTL, MSBB parahippocampal gyrus pQTL or MiGA microglial eQTL dataset. We use context for the tissue, brain region, cell type or cell subtype; molecular phenotype for the measured molecular layer, including gene expression, splicing, protein abundance, DNA methylation or histone acetylation; molecular target for the assayed gene, splice event, protein, CpG site or H3K9ac peak; and xQTL association for a variant-target association detected within the relevant analysis window. This manuscript describes the upstream FunGen-xQTL Atlas workflow and release. The atlas-generation workflow produces harmonized xQTL association files, significance-filtered files, fine-mapping outputs and portal-ready indexed datasets.

### Alzheimer’s Disease Sequencing Project (ADSP) Functional Genomics Consortium (FunGen-AD)

FunGen-AD was established through the National Institute on Aging (NIA) Functional Genomics Initiative (RFA-AG-21-006), launched in 2020 to bridge the gap between genetic variant identification and functional characterization in Alzheimer’s disease and related dementias (AD/ADRD). This initiative supports 12 distinct research grants across multiple institutions, each designed to elucidate the functional consequences of genetic variation and identify genetics-guided therapeutic targets for AD/ADRD prevention, diagnosis, and treatment. The xQTL Working Group within this consortium specifically focuses on generating a comprehensive reference map of QTLs that determine how genetic variation influences molecular traits across brain tissues and cell types relevant to AD pathogenesis. Full consortium membership and grant details are catalogued at https://adsp-fgc.niagads.org/research/.

### FunGen-xQTL Working Group

The xQTL Working Group coordinates all activities defined within the xQTL Project. Participating investigators contributed samples funded through their respective NIA grants, including cohorts from the Accelerating Medicines Partnership for Alzheimer’s Disease (AMP-AD), Knight-ADRC, and the MiGA. Only the MSBB and ROSMAP cohorts from the AMP-AD program were included in the current xQTL analyses (see **Supplementary Methods** for details on the analyzed cohorts). Complete working group membership, including contributions from Boston University, Icahn School of Medicine at Mount Sinai, University of Pennsylvania, and Washington University in St. Louis, is detailed in the ***Acknowledgements*** section.

### FunGen-AD Data Standardization Working Group

The FunGen-AD investigators, ADSP geneticists, and members of the NIAGADS^16^ established the FunGen-AD data standardization working group to develop data standards facilitating data collection, integration, and sharing. We used the same genome build (GRCh38/hg38) and reference gene models (ENSEMBL v103) as other ADSP sequencing data^37^. For downstream analyses that require LD information, the release is accompanied by a European-ancestry ADSP/GCAD LD reference panel using a stochastic genotype approach, available through NIAGADS^16,34^ (**Code Availability**).

### Data flow of the xQTL study

The Methods workgroup developed preprocessing and QC pipelines for sample phenotypes, genotypes, covariates and molecular phenotypes within each cohort; the full workflow is shown in **Supplementary Fig. S1**. Briefly, genotype data from sequencing and array platforms were imputed where required and subjected to uniform QC. Molecular phenotypes then underwent data type-specific QC, normalization, imputation for sparse assays and coordinate mapping. Association testing incorporated demographic covariates, genetic principal components and inferred hidden factors, and was performed with TensorQTL^38^ across cohorts and molecular phenotypes. Using the harmonized, QC-filtered inputs, we generated bulk and single-nucleus eQTLs, as well as splicing, protein, methylation and histone acetylation QTLs, for the datasets listed in **Table 1**. Each cohort applied the same pipelines to its own data. Finally, we harmonized xQTLs across cohorts and molecular phenotypes using hipFG^25^, producing searchable formats with shared variant and gene annotations and normalized effect statistics (**Code Availability**).

### Molecular phenotype preprocessing

After the common phenotype- and sample-level QC, we performed molecular phenotype-specific processing and QC (**Supplementary Methods**). We annotated each molecular phenotype with gene names and positions using ENSEMBL v103^39^, allowing conversion to BED format. We imputed missing phenotype values for all molecular phenotypes except bulk and single-nucleus RNA-seq data using gEBMF (implemented in the flashier package using empirical Bayes matrix factorization)^40^. Finally, we partitioned the molecular phenotype BED files by chromosome to ease processing/memory requirements.

### Constructing TAD-boundary-enhanced analysis regions

Brain cortex-specific and hippocampus-specific TADs were obtained from published datasets^41,42^. Within each tissue-specific dataset, TADs exhibited no internal overlap. The cortex and hippocampus TAD files were merged, with exactly duplicate TADs (identical chromosome, start, and end positions) removed. Three types of TAD overlaps were systematically resolved: (i) overlap between a TAD and its preceding TAD, (ii) overlap between a TAD and its following TAD, and (iii) a TAD entirely contained within another TAD. For type (i) overlaps, the start position of the overlapping TAD was extended to match the preceding TAD’s start position. Type (ii) overlaps were resolved by extending the end position to match the following TAD. Type (iii) contained TADs had both start and end positions extended to match their containing TAD. TADs that were completely contained within or exactly matched another TAD were removed. This iterative process of overlap identification, boundary extension, and TAD reduction was repeated until the TAD set reached a stable configuration.

Generalized TADs were constructed by identifying tissue TAD pairs exhibiting greater than 80% reciprocal overlap, which were combined into single generalized domains. TAD pairs not meeting this threshold were designated as sole-formed generalized TADs. TAD-boundary domains were assembled by defining and extending left (upstream) and right (downstream) borders. Left borders correspond to the start position of sole-formed generalized TADs or the first overlapping position of combined generalized TADs. Right borders were analogously defined using end positions. The complete list of generalized TAD-boundary regions served as the foundation for constructing gene-specific extended TAD-boundary regions. All genes within each TAD-boundary region were identified, and each gene’s transcription start and end positions were extended by 1 Mb to define the conventional gene-centered *cis* window. The boundaries of the gene’s occupied generalized TAD-boundary region and its 1 Mb *cis* window were combined, retaining the outermost boundaries to define the TAD-boundary-enhanced *cis* window. Genes residing within multiple generalized TAD-boundary regions incorporated boundaries from all relevant TAD-boundary regions in this calculation. Extremely large TAD-boundary regions were winsorized at the 99th percentile of genome-wide region length, corresponding to approximately 20 Mb, before downstream association testing.

### Calling significant xQTL associations

TensorQTL version 1.0.10 was executed with PLINK-format genotype files, BED-format phenotype files and covariate matrices as required inputs^38^. For eQTLs, snuc-eQTLs, pQTLs and sQTLs, TAD-boundary-enhanced *cis* regions were used as custom gene-based *cis* windows. Because of computational constraints, haQTLs and mQTLs used standard 1 Mb *cis* windows.

### Target-level Benjamini-Hochberg (BH) correction on xQTL associations

Following hipFG-based normalization and QC^25^, xQTLs were saved as separate file sets according to association significance. Target-level BH correction was carried out for each molecular target, such as a histone acetylation peak for haQTLs or a gene for eQTLs, using the multipletests function from statsmodels.stats.multitest with method set to “fdr_bh”. BH-significant associations were defined by BH FDR < 0.05, nominal p-value < 0.01 and minor allele samples > 1.

### Hierarchical multiple testing (HMT) correction on xQTL associations

HMT was carried out for each dataset according to Huang *et al.*, 2018^43^. Beginning with all tested variant-target associations, we calculated a Bonferroni-adjusted p-value for each QTL target as the product of the minimum nominal *p-*value and the number of variants tested for that target. These target-level adjusted *p*-values were then corrected by BH, and molecular targets with BH FDR < 0.05 were selected as significant targets. The maximum Bonferroni-adjusted *p*-value among the selected targets was used as the global threshold to filter HMT-significant associations.

### xQTL fine-mapping

We performed statistical fine-mapping of xQTL signals using the Sum of Single Effects (SuSiE) framework^28^. For expression, splicing, and protein QTLs, we applied SuSiE^28^ directly to individual-level genotype and phenotype data. For DNA methylation and histone acetylation QTLs, which consist of spatially correlated measurements at many CpGs or H3K9ac peaks within a region, we applied fSuSiE^29^, which extends SuSiE by modeling the spatial structure of genetic effects across nearby molecular locations through wavelet-based functional regression. Both methods decompose genetic effects into a sum of single-effect components, return a PIP for each variant, and produce 95% credible sets (CSs) designed to contain at least one causal variant with probability 0.95.

All input data followed the same QC procedures described above, and we used the default parameters of the susieR^28^ and fsusieR^29^ packages, with the maximum number of single-effect components initialized at 10 and increased as needed for regions with additional independent signals. fSuSiE used its default Daubechies least-asymmetric wavelets with 10 vanishing moments and an independent-shrinkage prior on the wavelet-transformed effects.

Across all molecular phenotypes, we retained CSs with a minimum absolute pairwise correlation of at least 0.8 among their variants. Within each retained CS, the sentinel variant was defined as the variant with the highest PIP, and CSs containing a single variant were defined as single-variant CSs, representing loci fine-mapped to variant-level resolution.

### hipFG-based annotation and harmonization

We standardized and quality-controlled xQTL and fine-mapping results using the hipFG harmonization pipeline^25^. This allowed us to standardize results into BED-like format, index them with Giggle for rapid querying^44^, remove records with invalid beta standard error statistics, remove records with unresolved alleles (expressed as asterisks), and ensure all alleles were left-aligned and effect-normalized. Allele verification and effect normalization enforced the non-reference allele as the effect allele, using dbSNP build 156^45^ and the GRCh38/hg38 reference genome. We annotated all variants and splicing, methylation, and histone acetylation targets to target genes and genomic features using ENSEMBL v103^39^. See **Supplementary Methods** for annotation details. All normalized tracks were annotated using the FILER metadata schema^46^.

### Within-atlas reproducibility analyses

To assess reproducibility within the atlas, we compared significant xQTL associations across datasets within the same QTL type when comparable targets were available. Associations were matched by variant and molecular target after hipFG harmonization^25^. For each dataset pair, effect-size concordance was summarized by the Spearman correlation of z-scores among shared associations. Signal sharing was summarized using the overlap coefficient, defined as the number of shared significant associations divided by the smaller number of significant associations in the two datasets. haQTLs were excluded from this within-atlas reproducibility analysis because only one haQTL dataset was available.

### Comparison with external eQTL resources

We obtained expression quantitative trait loci (eQTL) summary statistics from two external resources: the Genotype-Tissue Expression (GTEx) Project (v8)^3^, and MetaBrain^5^. To compare with our xQTL eQTL datasets, both GTEx and MetaBrain summary statistics were first normalized using hipFG. For GTEx, we used the significant eQTLs as defined by the GTEx project (FDR < 0.05). For MetaBrain, we used as significant all eQTLs with the per-target FDR < 0.05 (computed using hipFG). To validate our association findings, we then overlapped the sets of significant eQTLs (variant-gene pairs) across xQTL, GTEx and MetaBrain to assess degree of replication between detected associations across the studies.

### xQTL data portal

We developed the FunGen-xQTL data portal (https://xqtl.niagads.org) to enable interactive querying and visualization of harmonized xQTL datasets across cohorts, brain regions, cell types, and molecular phenotypes. The portal integrates expression, splicing, protein, methylation, and histone acetylation QTLs, together with fine-mapping results, from ROSMAP, MSBB, MiGA, and Knight-ADRC.

We harmonized all xQTL summary statistics using the hipFG framework^25^, which standardizes variant representation, allele orientation, genomic coordinates, and metadata across studies. We converted harmonized outputs into BED-like formats and indexed them with Giggle^44^ to enable rapid genomic interval queries. We stored indexed datasets in an AWS-based infrastructure to support scalable storage and low-latency retrieval.

We implemented query functionality for variant rsIDs or genomic coordinates, gene symbols and locus genomic-region coordinates. Queries were resolved using Giggle-based interval searches and returned as structured tables containing HMT associations and cross-cohort summaries.

We implemented genomic visualization using the NIAGADS GenomicsDB genome browser (IGV.js-based)^15^, which renders xQTL tracks in genomic context and supports visualization of fine-mapped credible sets and locus-level association patterns.

## Data Availability

We provide all xQTL summary statistics in tab-delimited formats (BED) through the NIAGADS Open Access Data Portal, with cross-referencing to the AD Knowledge Portal for integrated access. Bulk downloads will be available through the NIAGADS Data Sharing Service (DSS) (https://dss.niagads.org/open-access-data-portal), Accession Number: NG00184), including complete association results, per-target BH-corrected associations, HMT-corrected associations and fine-mapping result sets in annotation-friendly format. These data will also be available through the AD Knowledge Portal soon.

We obtained raw molecular phenotype data from the original cohort via NIAGADS (ng00083, ng00102, ng00114, ng00105) and the AD Knowledge Portal (syn20801188, syn21347564, syn21347197, syn31512863, syn52293417, syn3388564, syn22024496, syn4896408, syn3157275, syn17015098)^7,9,11,12,14–20^.

We obtained processed genotypes and companion datasets from NIAGADS (ng00067, ng00105, ng00127)^7,12,30^. Additional details for included datasets are provided in **Table 1**.

## Code Availability

All analysis code and processing pipelines are summarized below:

- End-to-end xQTL analysis framework (main repository): A modular, reproducible pipeline for multi-omic QTL analysis, encompassing genotype QC, molecular phenotype preprocessing for expression, splicing, protein, methylation and histone traits, reference data construction, including TAD-boundary and LD panels, QTL association testing, fine-mapping and harmonization: https://statfungen.github.io/xqtl-protocol/.
- xQTL analysis protocol and documentation: A comprehensive, web-based protocol describing the full xQTL analysis framework, including data preprocessing, reference data generation, TensorQTL association testing, fine-mapping and harmonization, with step-by-step guidance and links to executable workflows. The association-testing notebook is available here: https://statfungen.github.io/xqtl-protocol/qtl_association_testing.html.
- Reference data preparation for xQTL analyses: Notebooks implementing the reference data layer of the pipeline, including downloading and formatting genome and annotation resources, generating derived structural annotations such as TAD-boundary annotations, and constructing LD blocks for genome-wide fine-mapping of summary statistics, along with methods creating companion LD reference panels. Details are available here: https://statfungen.github.io/xqtl-protocol/reference_data.html.
- Specific workflows to highlight:

- Bulk RNA-seq expression preprocessing pipeline: A standardized workflow for processing bulk RNA-seq data, including read-level QC, alignment, gene-level quantification, filtering, and normalization to generate analysis-ready expression matrices for downstream eQTL mapping. Details are available here: https://statfungen.github.io/xqtl-protocol/bulk_expression.html.
- Alternative splicing QTL preprocessing pipeline: A workflow for quantifying and processing splicing variation from RNA-seq data, including intron clustering (e.g., via Leafcutter), filtering, normalization, and formatting into analysis-ready matrices for downstream sQTL mapping. Details are available here: https://statfungen.github.io/xqtl-protocol/splicing.html.
- snRNA-seq pseudobulk preprocessing pipeline: A standardized workflow for processing single-nucleus RNA-seq data, including cell-type stratification, pseudobulk aggregation, gene filtering, normalization, and batch correction to generate cell-type-specific expression matrices for downstream eQTL mapping. Details are available here: https://statfungen.github.io/xqtl-protocol/snRNAseq_preprocessing.html.
- DNA methylation QTL preprocessing and calling pipeline: A standardized workflow for processing Illumina 450K methylation data, including probe-level QC, normalization, transformation to beta and M values, missing data imputation, and formatting into analysis-ready matrices for downstream mQTL mapping. Details are available here: https://statfungen.github.io/xqtl-protocol/methylation_calling.html.
- H3K9ac QTL preprocessing and calling pipeline: A pipeline for processing H3K9ac ChIP-seq data to call and quantify acetylation peaks, producing molecular phenotype inputs for downstream QTL analyses. Details are available here: https://github.com/StatFunGen/xqtl-protocol/tree/main/code/script/molecular_phenotypes/h3k9ac_calling.
- Protein QTL processing and calling pipeline: A workflow for processing raw protein abundance data and imputing missing values to produce cleaned phenotype matrices suitable for downstream pQTL analysis. Details are available here: https://statfungen.github.io/xqtl-protocol/phenotype_imputation.html.
- TAD-boundary-enhanced *cis* region construction for xQTL mapping: A methodological workflow that constructs generalized TAD-boundary regions as biologically informed *cis* regions, integrating TAD data across cortex and hippocampus and systematically extending boundaries to capture regulatory variation beyond fixed windows. Details are available here: https://statfungen.github.io/xqtl-protocol/generalized_TADB.html.
- Univariate xQTL calling: A TensorQTL-based protocol for cis- and trans-xQTL mapping that takes genotype, phenotype and covariate inputs, supports standard and custom cis windows, and produces nominal, permutation-based and summary association outputs for downstream post-processing. Details are available here: https://statfungen.github.io/xqtl-protocol/qtl_association_testing.html.
- hipFG harmonization pipeline: https://bitbucket.org/wanglab-upenn/hipfg.

## Acknowledgements

We thank the xQTL Working Group members for their contributions to data generation, analysis, and harmonization: Columbia University (Xuewei Cao, Yanghyeon Cho, Natacha Comandante-Lou, Rui Dong, Masashi Fujita, Hans Klein, Yiyi Ma, Alexander McCreight, Min Qiao, Sheema Sameen, Badri Vardarajan, Anqi Wang, Lu Zeng); Boston University (Alexandre Pelletier, Pengjiao Wang, Mintao Lin, Oluwatosin Olayinka); Icahn School of Medicine at Mount Sinai (Alison Goate, Haochen Sun, Alan Renton); University of Pennsylvania (Travyse Edwards); Washington University in St. Louis (Chengran Yang). We gratefully acknowledge NIAGADS for data management and portal development, the FunGen-AD MPI members for scientific oversight, and the brain donors and their families for making this research possible.

This work was supported in part by NIH grants U54AG052427 and U24AG041689 (J.C., L.C., E.G.-A., Z.K., O.V., A.B.K., H.W., P.P.K., L.-S.W., and Y.Y.L.); U01AG072572 and R01AG076901 (R.F., P.L.D., and G.W.); R01AG076901 and The Urbut Family Foundation (A.L. and R.L.); U01AG072577, R01AG080810, and R01AG086467 (F.P.G., J.A.E., J.TCW., and X.Z.); U19AG068753, R01AG048927, U01AG062602, P30AG072878, U01AG082665, and U01AG081230 (L.A.F.); U01AG068880, R21AG063130, R01AG054005, RF1AG065926, and R01AG065926 (T.R.); R01AG93879, RF1AG077828, R21AG077168, and U01AG046170 (M.W.); and the Freedom Together Foundation (H.S.).

Whole-genome sequencing data for this study were processed and harmonized by the Genome Center for Alzheimer’s Disease (GCAD: U54AG052427) and prepared, archived, and distributed by the National Institute on Aging Alzheimer’s Disease Data Storage Site (NIAGADS: U24AG041689), both at the University of Pennsylvania and funded by the National Institute on Aging. The LD reference panel was developed by the FunGen-xQTL Project with support from NIH/NIA grants R01AG076901 and R01AG086467 (PI: Gao Wang), U01AG072572 (PI: Philip L. De Jager), and U01AG072577 (PI: Xiaoling Zhang). The panel was derived from ADSP Release 4 whole-genome sequencing data (European subset) obtained through NIAGADS.

ROSMAP: We are grateful to the participants in the Religious Orders Study and the Memory and Aging Project. This work is supported by NIH grants U01AG046152, R01AG043617, R01AG042210, R01AG036042, R01AG036836, R01AG032990, R01AG018023, RC2AG036547, P50AG016574, U01ES017155, KL2RR024151, K25AG041906, R01AG030146, P30AG010161, R01AG017917, R01AG015819, K08AG034290, and R01AG011101.

Mount Sinai Brain Bank (MSBB): This work was supported by NIH/NIA grants R01AG046170, RF1AG054014, RF1AG057440, and R01AG057907. R01AG046170 is a component of the AMP-AD Target Discovery and Preclinical Validation Project. Brain tissue collection and characterization was supported by NIH HHSN271201300031C.

## Author Contributions

P.P.K., X.Z., G.W., Y.Y.L., and led the project. J.C., R.F., F.P.G., L.C., H.C., P.P.K., G.W., and Y.Y.L. analyzed data. J.C., F.P.G., L.C., P.P.K., G.W., X.Z., and Y.Y.L. wrote the manuscript with input from all authors. R.F., F.P.G., A.L., H.S., R.L., J.A.E., J.TCW., M.W., and H.K. prepared data. E.G.-A., Z.K., O.V., A.B.K., and H.W. contributed infrastructure and data management. P.L.D., A.M.G., T.R., M.W., E.M., C.C., L.A.F., L.-S.W., X.Z., P.P.K., G.W., and Y.Y.L. contributed data, resources, and funding. The FunGen-AD Consortium contributed data and scientific input. All authors reviewed and approved the manuscript.

## References

1. Watanabe, K., et al. A global overview of pleiotropy and genetic architecture in complex traits. Nat Genet 51, 1339–1348 (2019).

2. Ng, B., et al. An xQTL map integrates the genetic architecture of the human brain’s transcriptome and epigenome. Nat Neurosci 20, 1418–1426 (2017).

3. Consortium, G.T. The GTEx Consortium atlas of genetic regulatory effects across human tissues. Science 369, 1318–1330 (2020).

4. Kerimov, N., et al. A compendium of uniformly processed human gene expression and splicing quantitative trait loci. Nat Genet 53, 1290–1299 (2021).

5. de Klein, N., et al. Brain expression quantitative trait locus and network analyses reveal downstream effects and putative drivers for brain-related diseases. Nat Genet 55, 377–388 (2023).

6. Jia, Y., et al. xQTLatlas: a comprehensive resource for human cellular-resolution multi-omics genetic regulatory landscape. Nucleic Acids Res 53, D1270–D1277 (2025).

7. Jang, B., et al. A meta-analysis of single-nucleus expression quantitative trait loci linking genetic risk to brain disorders. Nat Genet 58, 737–747 (2026).

8. Fernandez, M.V., et al. Genetic and multi-omic resources for Alzheimer disease and related dementia from the Knight Alzheimer Disease Research Center. Sci Data 11, 768 (2024).

9. Bellenguez, C., et al. New insights into the genetic etiology of Alzheimer’s disease and related dementias. Nat Genet 54, 412–436 (2022).

10. Yang, C., et al. Genomic atlas of the proteome from brain, CSF and plasma prioritizes proteins implicated in neurological disorders. Nat Neurosci 24, 1302–1312 (2021).

11. Sieberts, S.K., et al. Large eQTL meta-analysis reveals differing patterns between cerebral cortical and cerebellar brain regions. Sci Data 7, 340 (2020).

12. De Jager, P.L., et al. A multi-omic atlas of the human frontal cortex for aging and Alzheimer’s disease research. Sci Data 5, 180142 (2018).

13. Lopes, K.P., et al. Genetic analysis of the human microglial transcriptome across brain regions, aging and disease pathologies. Nat Genet 54, 4–17 (2022).

14. Lyon, M.S., et al. The variant call format provides efficient and robust storage of GWAS summary statistics. Genome Biol 22, 32 (2021).

15. Greenfest-Allen, E., et al. NIAGADS Alzheimer’s GenomicsDB: A resource for exploring Alzheimer’s disease genetic and genomic knowledge. Alzheimers Dement 20, 1123–1136 (2024).

16. Kuzma, A., et al. NIAGADS: A data repository for Alzheimer’s disease and related dementia genomics. Alzheimers Dement 21, e70255 (2025).

17. Greenwood, A.K., et al. The AD Knowledge Portal: A Repository for Multi-Omic Data on Alzheimer’s Disease and Aging. Curr Protoc Hum Genet 108, e105 (2020).

18. Mathys, H., et al. Single-cell atlas reveals correlates of high cognitive function, dementia, and resilience to Alzheimer’s disease pathology. Cell 186, 4365–4385 e4327 (2023).

19. Fujita, M., et al. Cell subtype-specific effects of genetic variation in the Alzheimer’s disease brain. Nat Genet 56, 605–614 (2024).

20. Mathys, H., et al. Single-cell multiregion dissection of Alzheimer’s disease. Nature 632, 858–868 (2024).

21. Comandante-Lou, N., et al. PLXNB1 and other signaling drives a pathologic astrocyte state contributing to cognitive decline in Alzheimer’s Disease. bioRxiv (2025).

22. Wang, M., et al. The Mount Sinai cohort of large-scale genomic, transcriptomic and proteomic data in Alzheimer’s disease. Sci Data 5, 180185 (2018).

23. Coleman, C., et al. Multi-omic atlas of the parahippocampal gyrus in Alzheimer’s disease. Sci Data 10, 602 (2023).

24. Dube, U., et al. An atlas of cortical circular RNA expression in Alzheimer disease brains demonstrates clinical and pathological associations. Nat Neurosci 22, 1903–1912 (2019).

25. Cifello, J., et al. hipFG: high-throughput harmonization and integration pipeline for functional genomics data. Bioinformatics 39 (2023).

26. Orchard, P., et al. Cross-cohort analysis of expression and splicing quantitative trait loci in TOPMed. medRxiv (2025).

27. Dixon, J.R., Gorkin, D.U. & Ren, B. Chromatin Domains: The Unit of Chromosome Organization. Mol Cell 62, 668–680 (2016).

28. Wang, G., Sarkar, A., Carbonetto, P. & Stephens, M. A simple new approach to variable selection in regression, with application to genetic fine mapping. J R Stat Soc Series B Stat Methodol 82, 1273–1300 (2020).

29. Denault, W.R.P., et al. fSuSiE enables fine-mapping of QTLs from genome-scale molecular profiles. bioRxiv (2025).

30. Chung, J., et al. Genome-wide association and multi-omics studies identify MGMT as a novel risk gene for Alzheimer’s disease among women. Alzheimers Dement 19, 896–908 (2023).

31. Hemming, M.L. & Selkoe, D.J. Amyloid beta-protein is degraded by cellular angiotensin-converting enzyme (ACE) and elevated by an ACE inhibitor. J Biol Chem 280, 37644–37650 (2005).

32. Oba, R., et al. The N-terminal active centre of human angiotensin-converting enzyme degrades Alzheimer amyloid beta-peptide. Eur J Neurosci 21, 733–740 (2005).

33. Messenger, E.J., et al. PLCG2 modulates TREM2 expression and signaling in response to Alzheimer’s disease pathology. Alzheimers Dement 21, e70231 (2025).

34. Leung, Y.Y., et al. Alzheimer’s Disease Sequencing Project release 4 whole genome sequencing dataset. Alzheimers Dement 21, e70237 (2025).

35. Wang, N., et al. StocSum: stochastic summary statistics for whole genome sequencing studies. bioRxiv (2023).

36. Chunming Liu, A.W., Hao Sun, Kaixuan Luo, Sheng Qian, Yining Li, Xin He, Phillip De Jager, David A Bennett, Minghui Wang, Carlos Cruchaga, The Alzheimer’s Disease Functional Genomics Consortium, Gao Wang, Fabio Morgante. A Multi-Context Regulome-Wide Association Atlas for Genetic Studies of Aging Brain Disorders. medRxiv (2026).

37. Leung, Y.Y., et al. VCPA: genomic variant calling pipeline and data management tool for Alzheimer’s Disease Sequencing Project. Bioinformatics 35, 1768–1770 (2019).

38. Taylor-Weiner, A., et al. Scaling computational genomics to millions of individuals with GPUs. Genome Biol 20, 228 (2019).

39. Harrison, P.W., et al. Ensembl 2024. Nucleic Acids Res 52, D891–D899 (2024).

40. Wang, W. & Stephens, M. Empirical Bayes Matrix Factorization. J Mach Learn Res 22(2021).

41. McArthur, E. & Capra, J.A. Topologically associating domain boundaries that are stable across diverse cell types are evolutionarily constrained and enriched for heritability. Am J Hum Genet 108, 269–283 (2021).

42. Schmitt, A.D., et al. A Compendium of Chromatin Contact Maps Reveals Spatially Active Regions in the Human Genome. Cell Rep 17, 2042–2059 (2016).

43. Huang, Q.Q., Ritchie, S.C., Brozynska, M. & Inouye, M. Power, false discovery rate and Winner’s Curse in eQTL studies. Nucleic Acids Res 46, e133 (2018).

44. Layer, R.M., et al. GIGGLE: a search engine for large-scale integrated genome analysis. Nat Methods 15, 123–126 (2018).

45. Sherry, S.T., et al. dbSNP: the NCBI database of genetic variation. Nucleic Acids Res 29, 308–311 (2001).

46. Kuksa, P.P., et al. FILER: a framework for harmonizing and querying large-scale functional genomics knowledge. NAR Genom Bioinform 4, lqab123 (2022).

