## Supplementary Methods for "A TAD-informed aging-brain xQTL atlas of multi-modal and cell-type-resolved regulatory variation"

### Supplementary Information

### Supplementary Notes

#### Supplementary Note 1. Processing, QC and harmonization workflow

The FunGen-xQTL Atlas data generation, harmonization and dissemination framework is summarized in **Fig. 1A** and **Supplementary** **Fig. S1**. This workflow begins with cohort-level genotype, molecular phenotype and phenotype inputs, proceeds through standardized preprocessing and quality control, and ends with harmonized association files, fine-mapping outputs and portal-ready indexed datasets.

The workflow was designed to make datasets generated by different contributing groups comparable at the level needed for reuse. Genotype processing aligned variant identifiers, alleles, coordinates and ancestry covariates where required. Molecular phenotypes were processed with data-type-specific pipelines for expression, splicing, protein abundance, DNA methylation and histone acetylation, followed by filtering, normalization, covariate construction and hidden-factor adjustment.

Association testing used a shared modeling framework across QTL types while allowing data-type-specific analysis windows and computational constraints. Expression, splicing and protein QTLs were analyzed with gene- or target-centered windows that could incorporate TAD-boundary-enhanced regions, whereas methylation and histone acetylation QTLs used standard distance-based windows.

After association testing and fine-mapping, outputs were passed through hipFG-based^1^ harmonization to standardize allele orientation, effect-size representation, genomic coordinates, target identifiers and metadata. The same framework produces the downloadable files, browser tracks and portal indices used in the public release, making the computational workflow itself part of the reusable resource.

#### Supplementary Note 2. Regional fine-mapping details at AD risk loci

At the *ACE* locus, fine-mapping resolved the BH-significant signals into eleven credible sets comprising thirty-three variants across five QTL types and five cohorts. Several splicing and methylation signals showed high resolution and mapped to distinct genomic positions. Two ROSMAP sQTLs localized to separate variants, including rs1444621153 (PIP = 1.00; 1.2 Mb from target) and rs547114353 (PIP = 0.96; +524 kb), each associated with different splice clusters. The Knight-ADRC mQTL resolved to a singleton at rs35543459 (PIP = 1.00; +56 kb). These signals did not overlap, indicating independent regulatory effects across splicing and methylation.

Expression-related signals at *ACE* were less resolved but showed concordance across datasets. The ROSMAP DLPFC eQTL produced a five-variant credible set led by rs4292 (PIP = 0.58; z = -11.6; -81 bp). This region was also supported by the MSBB pQTL, where rs4292 was the top variant in a six-variant credible set (PIP = 0.38), and by two single-nucleus excitatory neuron eQTL datasets, where rs4291 led nine-variant credible sets (PIP = 0.28 and 0.26). These four signals showed consistent negative effect directions (z-scores from -5.1 to -11.6), supporting a shared regulatory component distinct from the splicing and methylation signals.

At the *PLCG2* locus, fine-mapping identified fourteen credible sets across four cohorts, eleven of which resolved to single variants. The strongest signal was observed for methylation, where rs7199820 reached PIP = 1.00 across three proximal CpG targets in ROSMAP DLPFC, with large negative effects (z-scores from -22 to -30) within 300 bp. Two additional singleton credible sets in Knight-ADRC provided independent support.

Expression signals at *PLCG2* showed greater allelic complexity. In ROSMAP DLPFC, three eQTL credible sets were identified, including a singleton (PIP = 1.00), a two-variant set (lead PIP = 0.74), and a seven-variant set (lead PIP = 0.27), consistent with multiple independent *cis* signals. Additional singleton eQTLs in ROSMAP PCC and MSBB frontal cortex, Brodmann area 22 supported this pattern. The variant rs8043593 reached PIP = 1.00 in both ROSMAP AC eQTL and sQTL analyses but had opposite effect directions (z = +39.7 for eQTL and z = -35.8 for sQTL), indicating distinct regulatory effects on expression level and splicing. The oligodendrocyte single-nucleus eQTL signal remained unresolved, spanning ten variants across 160 kb with a maximum PIP of 0.30.

### Supplementary Methods

#### Molecular dataset composition

The FunGen-xQTL Atlas integrates molecular datasets from ROSMAP, MSBB, Knight-ADRC and MiGA. We use molecular dataset for a cohort- or source-specific molecular measurement panel used for xQTL mapping, context for the tissue, brain region, cell type or cell subtype, modality for the measured molecular layer, and molecular target for the assayed gene, splice event, protein, CpG or H3K9ac peak. The complete dataset-level inventory and sample sizes are provided in **Table 1** and **Supplementary Tables S2** and **S3**.

ROSMAP contributed bulk RNA-seq eQTL datasets from DLPFC, PCC and AC, bulk sQTL datasets from the same three regions, and DLPFC pQTL, mQTL and H3K9ac haQTL datasets. It also contributed peripheral blood monocyte eQTLs and multiple DLPFC single-nucleus RNA-seq resources, including CUIMC1, MIT and a CUIMC1/CUIMC2/MIT mega-analysis. The CUIMC1 resource contains six major brain cell types, astrocytes, excitatory neurons, inhibitory neurons, microglia, oligodendrocytes and oligodendrocyte progenitor cells, with 418 to 419 samples per cell type. The MIT resource contains the same six major cell types with 377 to 387 samples per cell type. The CUIMC1/CUIMC2/MIT mega-analysis contains the same six major cell types with 733 to 737 samples per cell type. CUIMC2 is a recently generated ROSMAP DLPFC single-nucleus multiome dataset whose RNA profiles from approximately 240 donors contributed to the mega-analysis rather than being released as a separate xQTL dataset in this atlas.

MSBB contributed bulk RNA-seq eQTL datasets from frontal pole (Brodmann area 10), superior temporal gyrus (Brodmann area 22), parahippocampal gyrus (Brodmann area 36) and inferior frontal gyrus (Brodmann area 44), with 230 to 274 samples per region, plus parahippocampal gyrus pQTL and mQTL datasets. Knight-ADRC contributed parietal cortex eQTL, pQTL and mQTL datasets. MiGA contributed microglial eQTL datasets from medial frontal gyrus, superior temporal gyrus, subventricular zone and thalamus, with 47 to 66 samples per region. Together, these panels define the molecular and cellular inputs used for FunGen-xQTL Atlas association testing, fine-mapping and release harmonization.

#### Cohort data summary

##### Knight-ADRC

The Charles F. and Joanne Knight Alzheimer’s Disease Research Center (Knight-ADRC) Memory and Aging Project cohort at Washington University School of Medicine in St. Louis enrolled individuals aged 65 years and older who demonstrated either no memory impairment or mild dementia at study entry^2^. Participants underwent comprehensive longitudinal cognitive assessments and were followed until death. At death, autopsied brains underwent standardized neuropathological evaluation using CERAD criteria for neuritic plaques and Braak staging for neurofibrillary tangles, with final diagnoses assigned according to NIA-AA guidelines. Molecular phenotype data collection focused on parietal lobe cortex tissue, with genetic, proteomic, transcriptomic and DNA methylation profiling performed on postmortem samples collected according to established Knight-ADRC autopsy protocols (<https://dss.niagads.org/collections/knight-adrc-collection>).

##### MSBB

The Mount Sinai/JJ Peters VA Medical Center Brain Bank (MSBB) cohort comprises postmortem brain tissue from individuals representing the full spectrum of Alzheimer’s disease (AD) severity, from preclinical to advanced stages. Cohort assembly followed a rigorous filtering protocol that included comprehensive neuropathological assessment of amyloid-beta plaques, neurofibrillary tangles, Lewy bodies and vascular pathology^3^. The minimum age at death for inclusion was 61 years. Molecular profiling targeted four anatomically defined Brodmann areas, frontal pole (BA10), inferior frontal gyrus (BA44), parahippocampal gyrus (BA36) and superior temporal gyrus (BA22). Tissue collection protocols standardized dissection procedures across brain regions to ensure consistent sampling from homologous cortical laminae for gene expression, protein expression and DNA methylation analyses. Cohort details are available through the AD Knowledge Portal (<https://www.synapse.org/Synapse:syn3159438>).

##### MiGA

The Microglia Genomic Atlas (MiGA) cohort was constructed from postmortem human brain samples obtained from the Netherlands Brain Bank (NBB) and the Neuropathology Brain Bank and Research CoRE at Mount Sinai Hospital, New York^4^. The cohort encompasses subjects with neurodegenerative diseases, other neurological/neuropsychiatric disorders, and neurologically normal controls. Primary microglia were isolated ex vivo from four distinct brain regions: medial frontal gyrus (MFG), superior temporal gyrus (STG), subventricular zone (SVZ), and thalamus (THA). Study participants exhibited a mean age at death of 73.6 years with an age range spanning 21 to 103 years. Microglia isolation protocols optimized yield and purity through immunomagnetic separation and fluorescence-activated cell sorting, preserving transcriptional profiles representative of in situ microglial states ([https://dss.niagads.org/sample-sets/snd10022](https://dss.niagads.org/sample-sets/snd10022/)).

##### ROSMAP

The Religious Orders Study (ROS) and Rush Memory and Aging Project (MAP), collectively known as ROSMAP, comprise older individuals without known dementia at enrollment who underwent annual standardized clinical assessments of global cognitive function and domain-specific cognitive performance^5^. Participants were assigned final cognitive diagnoses at death according to established criteria: no cognitive impairment, mild cognitive impairment (MCI), or syndromic AD. The cohort demonstrates a mean age at death of 88.8 years. Molecular phenotype profiling utilized gray matter dissected from the dorsolateral prefrontal cortex (DLPFC) and anterior caudate (AC). Longitudinal clinical data enabled correlation of molecular quantitative trait loci (QTL) findings with antemortem cognitive trajectories and neuropathological outcomes assessed through standardized autopsy protocols. <https://adknowledgeportal.synapse.org/Explore/Studies/DetailsPage/StudyDetails?Study=syn3219045>

#### Genotype data processing

##### Array imputation protocols

For the MiGA cohort, genotype imputation was performed using the Michigan Imputation Server v1.4.1 with the 1000 Genomes Project Phase 3 version 5 (GRCh37) European reference panel and Eagle version 2.4 for phasing^6-8^. Following imputation, variants were lifted over to the GRCh38 reference to match the RNA-seq data using Picard LiftoverVcf and the b37ToHg38.over.chain.gz liftover chain file. The Knight-ADRC cohort genotypes were imputed using the TOPMed Imputation Server with the TOPMed reference panel release r2, as detailed in the original publication^9^. Imputation quality control followed the protocols established in each cohort’s original study.

##### WGS variant calling thresholds

Whole genome sequencing data underwent variant calling quality control using bcftools version 1.21^10^. Multi-allelic sites were split into bi-allelic records and annotated using dbSNP build 156^11^. Allelic balance for heterozygous genotypes was calculated as the number of bases supporting the less-represented allele divided by the total number of base observations. VCF files were converted to PLINK format version 1.90b7.7, with maximum missingness of 0.1 enforced for both variants and samples^12^.

##### Relatedness and PCA-based outlier removal

Principal components were calculated using unrelated individuals in each cohort to be used as covariates in QTL calling and fine-mapping. KING version 2.3.0 was used to identify the maximum set of unrelated individuals, with related individuals removed based on kinship coefficients of 0.25 (first-degree), 0.125 (second-degree), and 0.0625 (third-degree), respectively^13^. Variant pruning was performed on the unrelated individual set using a linkage disequilibrium threshold of r² > 0.1. Principal component analysis (PCA) was conducted on unrelated samples with less than 10% variant missingness, and statistical outlier individuals were removed based on principal component (PC) deviation. Individuals were projected into the resulting PC space.

#### Sample phenotype preprocessing

Sex, age at death and post-mortem interval (PMI) were recorded for all samples, except MiGA, for which PMI data were unavailable. ROSMAP includes study origin (ROS or MAP) as a covariate, and MSBB includes RNA integrity number (RIN). In addition to fixed covariates, we calculated genetic PCs for all cohorts and retained PCs explaining 70% of variance. We then performed PCA-based hidden-factor analysis on molecular phenotype data for each sample, with automatic determination of the number of factors to retain^14^. A sample-covariate matrix was generated with these components and included in TensorQTL xQTL calling (**Methods**, Calling significant xQTL associations).

#### Molecular phenotype data processing

##### Methylation array specifications

Methylation data from the Illumina Infinium HumanMethylation450 BeadChip arrays were processed from IDAT files using the Sesame software package version 1.0.3^15^. Processing included poor-probe masking (Q), channel inference for Infinium-I probes (C), non-linear dye-bias correction (D), detection p-value masking using out-of-band probes (P), and out-of-band background subtraction (B). M-values were calculated as M = log2(beta/(1-beta)), where beta = 0 or beta = 1 values were replaced by the next minimum or maximum values in the beta matrix, respectively. Samples with less than 80% probe detection were excluded. Probes with greater than 5% missingness across samples were removed. Remaining missing values in the logit-transformed beta matrix were imputed using flashier, an empirical Bayes matrix factorization method.

##### Bulk RNA-seq processing

Raw FASTQ reads underwent quality assessment with fastqc version 0.12.1 followed by adapter trimming using fastp version 0.24.1^16^. Reads were aligned to the GRCh38 reference genome using STAR version 2.7.11b with the following parameters^17^:

--outSAMtype BAM SortedByCoordinate --outFilterMultimapNmax 20 --outFilterMismatchNmax 999 --outFilterMismatchNoverLmax 0.04 --alignIntronMin 20 --alignIntronMax 1000000 --alignMatesGapMax 100000 --outFilterMatchNminOverLread 0.95 --outSJfilterReads Unique --outSJfilterCountUniqueMin 3 --outSJfilterCountTotalMin 5 --outSJfilterDistMean 1000 --outSJfilterDistStdDevScaled 5 --chimSegmentMin 10 --chimOutType WithinBAM SoftClip --chimJunctionOverhangMin 10 --chimScoreDropMax 30 --chimScoreJunctionNonGTAG 0 --chimScoreSeparation 1 --chimSegmentReadGapMax 3 --chimMultimapNmax 20.

Gene-level expression was quantified using RNA-SeQC version 2.4.2 and transcript-level expression using RSEM version 1.3.3 with the GRCh38.103 Ensembl GTF reference^18^.

##### Splicing quantification

Alternative splicing events were quantified using LeafCutter2^19^, which identifies intron excision usage, skipped exons and alternative splice site usage. Unlike gene-expression quantification, splicing analysis used BAM files generated with the WASP option in STAR^17^ to correct for allele-specific expression bias. LeafCutter2^19^ output was converted to BED format. Splicing features missing in more than 40% of samples, introns with variation less than 0.001, and events flagged as inconclusive or not expressed were excluded before downstream analysis. Quantile-quantile normalization was applied, followed by imputation using empirical Bayes matrix factorization. Splicing features were then mapped to genes using ENSEMBL GRCh38.103 annotations.

##### Single-nucleus RNA-seq processing

Single-nucleus RNA-seq data were obtained as published R objects generated using Seurat version 5.2.1^20^. Seurat objects were subset by cell type to create separate objects for astrocytes, inhibitory neurons, oligodendrocytes, oligodendrocyte progenitor cells and microglia^21,22^. The excitatory-neuron Seurat object was generated by combining three excitatory-neuron subtype files from Green et al. 2024^22^, retaining only genes shared across all three matrices and summing expression values for samples appearing in multiple files. Pseudobulk profiles were generated by aggregating raw count data per sample using the Seurat AggregateExpression function, with all samples containing fewer than 10 cells removed. Genes were filtered using filterByExpr to retain features with sufficient expression across samples. Trimmed mean of M-values (TMM) normalization was applied to adjust for library composition effects, followed by transformation to log2-counts per million (logCPM), with genes exhibiting mean logCPM < 2.0 removed. Final quantile normalization ensured consistent expression distributions across samples. Before constructing the CUIMC1/CUIMC2/MIT mega-analysis, overlapping ROSMAP donors were resolved by retaining CUIMC1 profiles when they overlapped with MIT or CUIMC2 and retaining MIT profiles when they overlapped with CUIMC2; CUIMC2 RNA profiles contributed for remaining non-overlapping donors. For the CUIMC1/CUIMC2/MIT mega-analysis, batch effects among the MIT and CUIMC datasets were removed using the limma removeBatchEffects function after gene-expression filtering, specifying MIT, CUIMC1 and CUIMC2 as separate batches^23^.

##### Proteomic data processing

Knight-ADRC protein data were generated using the SomaScan aptamer-based platform, measuring the relative abundance of 1,305 proteins from parietal lobe cortex samples of 458 individuals. Sample- and aptamer-level quality control was performed as described^2,24^. MSBB Brodmann area 36 samples were analyzed by tandem mass tag mass spectrometry (TMT-MS). Proteins with missingness exceeding 15% were removed, resulting in 2,962 proteins retained. Protein abundance values were adjusted using a mixed-effects model incorporating a random effect term for batch correction^25^. ROSMAP DLPFC and temporal cortex/precuneus samples from 453 individuals underwent TMT-MS analysis using procedures matching those used for MSBB. Detected peptides were filtered by false discovery rate and assembled into 5,688 unique proteins, with a final 50% missingness filter yielding 3,334 proteins^26^.

##### H3K9ac ChIP-seq peak calling

Histone H3 lysine 9 acetylation (H3K9ac) peaks were called from Millipore anti-H3K9ac antibody ChIP-seq experiments performed on dorsolateral prefrontal cortex samples from the ROSMAP cohort^27^. Peaks were identified using the MACS2 callpeak function with the --broad option, q-value cutoff of 0.05, and broad cutoff of 0.1^28^. Quality control metrics for MACS2 outputs and intermediate files were compiled using MultiQC, and called peak regions were overlapped with the Roadmap Epigenomics 15-state chromatin model to characterize relative chromatin state frequencies^29^. Custom scripts using the GenomicRanges R package processed peak output files to calculate peak ranges and define peak domains^30^. H3K9ac probes were annotated using genomic position for each peak ID, and peaks overlapping ENCODE blacklist regions were excluded from downstream analysis^31^. Finally, H3K9ac peak counts were TMM-normalized and fit to experimental covariates using the lmFit and eBayes functions from the R limma package. The residuals from this fitting procedure were used as H3K9ac expression values.

#### Summary statistics genomic feature annotation

##### Variant, haQTL-, and mQTL-target annotation

All variants and targets for haQTLs and mQTLs were annotated using two genomic feature partitions constructed from ENSEMBL v103^18^. The first ("coding") partition was created by keeping only protein coding, processed pseudogene, and TR and IG gene biotypes. Within each transcript, introns were added between exonic elements (five_prime_utr, CDS, three_prime_utr, start_codon, and stop_codon) to create full continuous transcripts. Promoters were added, defined as the 10 kb upstream of the gene transcription start site (TSS). The second ("noncoding") partition filtered for non-protein coding gene biotypes and preserved exon segments, with introns again added in between to create a full transcript.

The partitions were queried using variant start positions, methylation target cytosine positions, and histone acetylation peak midpoint positions. The resulting overlaps used overlapping genes when possible and mapped according to the nearest gene TSS when not. In the case of multiple gene overlaps, genomic feature types were used to prioritize, in descending order of priority, coding exons, 5′ UTR exons, 5′ UTR introns, 3′ UTR exons, 3′ UTR introns, coding introns, noncoding exons, noncoding introns, promoters, and intergenic regions. If the only coding overlap was a promoter, the noncoding overlaps were used instead. Variants had all overlaps reported and ha/mQTL targets had the highest-priority mapping assigned to the target_gene_symbol and target_ensembl_id fields.

##### sQTL target annotation

For splicing targets, we assigned target genes based on stranded overlaps between splicing regions and gene annotations. We overlapped splicing regions with the ENSEMBL v103^18^ gene annotations (without the above gene-biotype or feature filtering). For each reported LeafCutter2^19^ cluster (span), we prioritized gene fragments with endpoints exactly matching splicing span, followed by those exactly matching one endpoint of the span, and lastly prioritizing fragments overlapping one span’s endpoint within a 10bp window. Following this overlap prioritization, two special cases allowed for two genes to be selected as annotations for spliced fragments: 1) both endpoints of the span exactly matched the exon endpoints for different genes (gene fusion) or 2) one endpoint was an exact match and the other was intragenic. For double intragenic overlaps, we selected the gene with a greater start position. Lastly, when only one gene was prioritized, we selected it for sQTL target annotation. Clusters with a strand assignment of "?" and cluster spans that failed to overlap any genes used their original LeafCutter2 expanded cluster annotations.

**Supplementary Tables**

**Supplementary Table S1.** Comparison of brain xQTL resources. Summary of the FunGen-xQTL Atlas relative to existing xQTL and QTL resources, including xQTL Serve^32^, GTEx v8^33^, MetaBrain^34^, the eQTL Catalogue^35^, Ontime browser^9^, and xQTL Atlas^36^. Resources are compared across reference, total number of brain QTL associations, association scope, availability of statistical fine-mapping and credible sets, assay types, brain regions, brain cell types, sample size, data harmonization and indexing, and availability of an interactive web portal for cross-QTL search and download. The FunGen-xQTL Atlas provides a harmonized, indexed, multi-assay brain xQTL resource spanning *cis* (±1Mbp) and TAD-defined (>1Mbp) regulatory domains with fine-mapped credible sets and interactive access through the NIAGADS xQTL portal.

**Supplementary Table S2.** Summary of xQTL association and fine-mapping counts across datasets. Counts of tested xQTL associations, Benjamini-Hochberg FDR-significant associations, HMT associations, and 95% credible set fine-mapping associations are shown for each dataset. Rows correspond to individual cohort, molecular phenotype, tissue, brain region, or cell-type contexts, including bulk eQTL, mQTL, pQTL, sQTL, haQTL, and single-nucleus eQTL datasets. This table summarizes the scale of association testing and the progressive refinement of signals from all tested variant–target pairs to significant associations and fine-mapped credible set associations.

**Supplementary Table S3.** Summary of *cis* (±1Mbp) and TAD-defined (>1Mbp) xQTL associations across datasets. Counts of tested associations, Benjamini-Hochberg FDR-significant associations, and variant categories are shown for each dataset using conventional *cis* windows and TAD-defined analysis windows. TAD association percentages indicate the proportion of associations identified within TAD-defined regions relative to the combined *cis* and TAD search space. Variant categories distinguish variants detected only in *cis* windows, only in TAD windows, or in both. Four datasets did not use TAD-defined windows and therefore have TAD counts reported as 0/NA: Knight-ADRC.mQTL.PC, MSBB.mQTL.PHG, ROSMAP.haQTL.DLPFC, and ROSMAP.mQTL.DLPFC.

**Supplementary Table S4**. All 206 high-confidence causal genes identified from 95% credible sets, using high-confidence (PIP > 0.8) variants. High-confidence genes were identified as appearing (in 95% credible sets) in at least 2 independent cohorts, at least 2 QTL types, at least 2 tissues or cell types, and having at least one credible set with more than one variant. Additionally, all genes are required to be targeted by at least one PIP>0.8 variant appearing in multiple QTL types, cohorts, and tissue or cell types. To remove complicated loci, genes targeted by more than 5 variants were removed. The cohorts ROSMAP, CUIMC1, MIT, and CUIMC1,2+MIT were considered not independent from each other for the purpose of counting independent cohorts.

**Supplementary Figures**

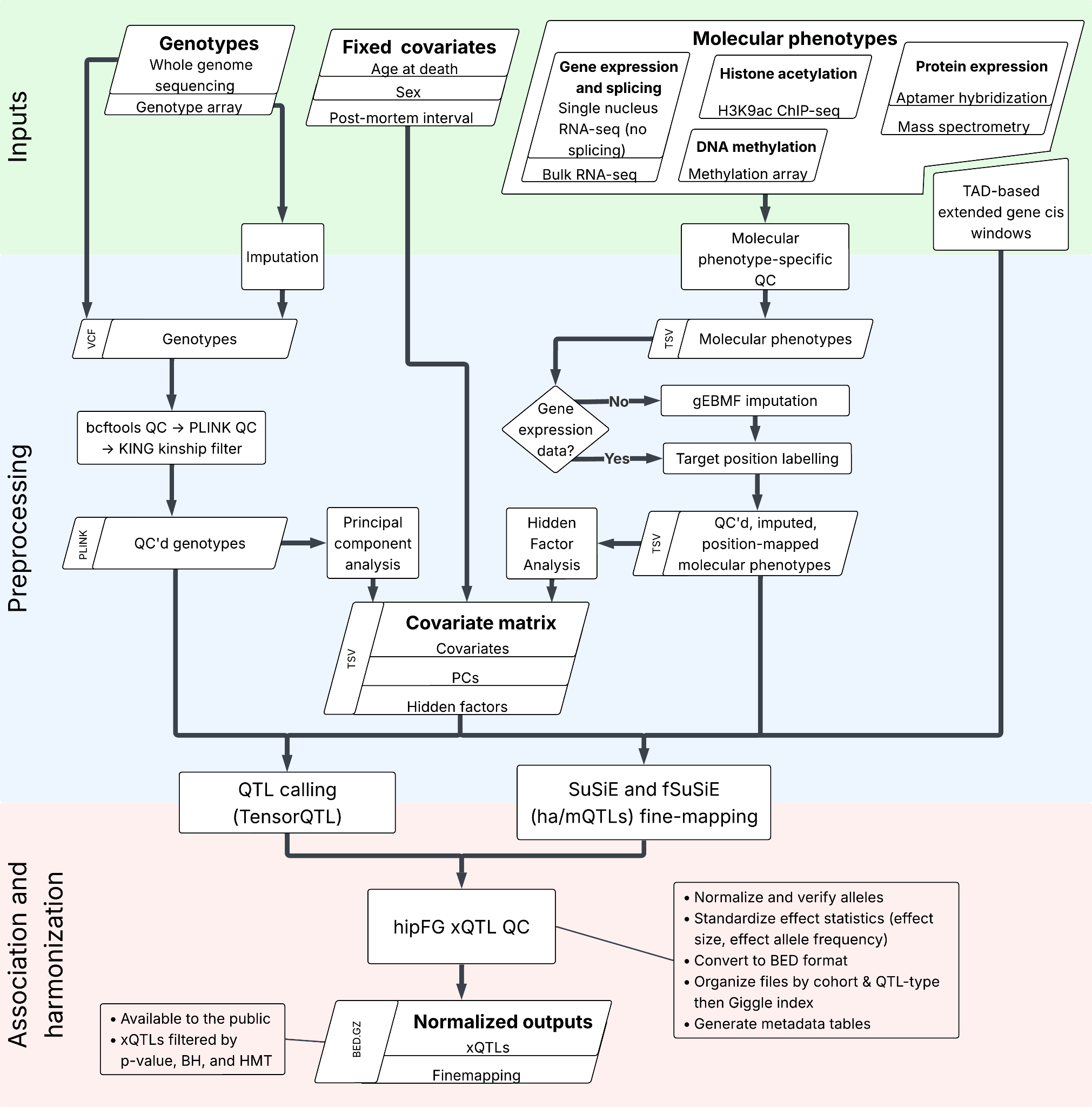

**Supplementary Fig. S1. Standardized xQTL data processing, association testing and harmonization framework.**

Schematic of the end-to-end computational workflow for xQTL analysis across cohorts and molecular phenotypes. Inputs include genotype data (whole genome sequencing (WGS) or genotyping arrays), fixed covariates (age at death, sex, and post-mortem interval) and molecular phenotypes encompassing gene expression (bulk and snuc RNA-seq) and splicing (bulk RNA-seq), histone acetylation (H3K9ac ChIP–seq), DNA methylation (array-based) and protein abundance (aptamer-based or mass spectrometry). Genotypes undergo imputation followed by quality control (bcftools and PLINK^12^), after which principal components are computed on unrelated individuals as determined by kinship analysis (KING)^13^. Molecular phenotypes are subjected to data type-dependent quality control including gEBMF imputation for non-expression molecular traits and mapping of all features to genomic coordinates. Molecular phenotype covariates are inferred via hidden factor analysis (**Supplementary Methods**). A unified covariate matrix is thus constructed from the fixed, genetic, and phenotypic covariates. xQTL mapping is performed using TensorQTL^33^ and fine-mapping is performed using SuSiE^37^ and fSuSiE^38^. Resulting summary statistics undergo harmonization using hipFG^1^, including allele normalization, effect size standardization, format conversion, and metadata generation, yielding uniformly formatted, indexed and publicly accessible xQTL and fine-mapping outputs (**Methods**).

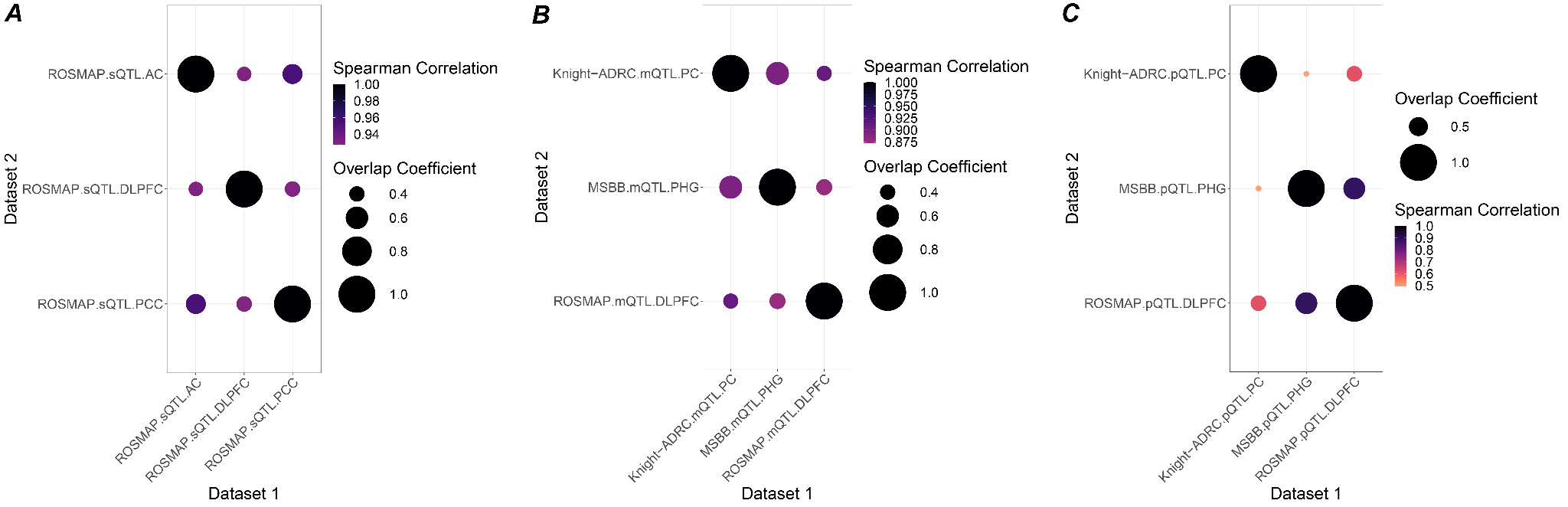

**Supplementary Fig. S2.** Replication plots across FunGen HMT-significant QTL types including **A)** sQTLs, **B)** mQTLs, and **C)** pQTLs. Spearman correlations are calculated based on z-scores of overlapping associations between two datasets. Overlap coefficient is calculated based on association overlap count divided by the association count of smaller dataset.

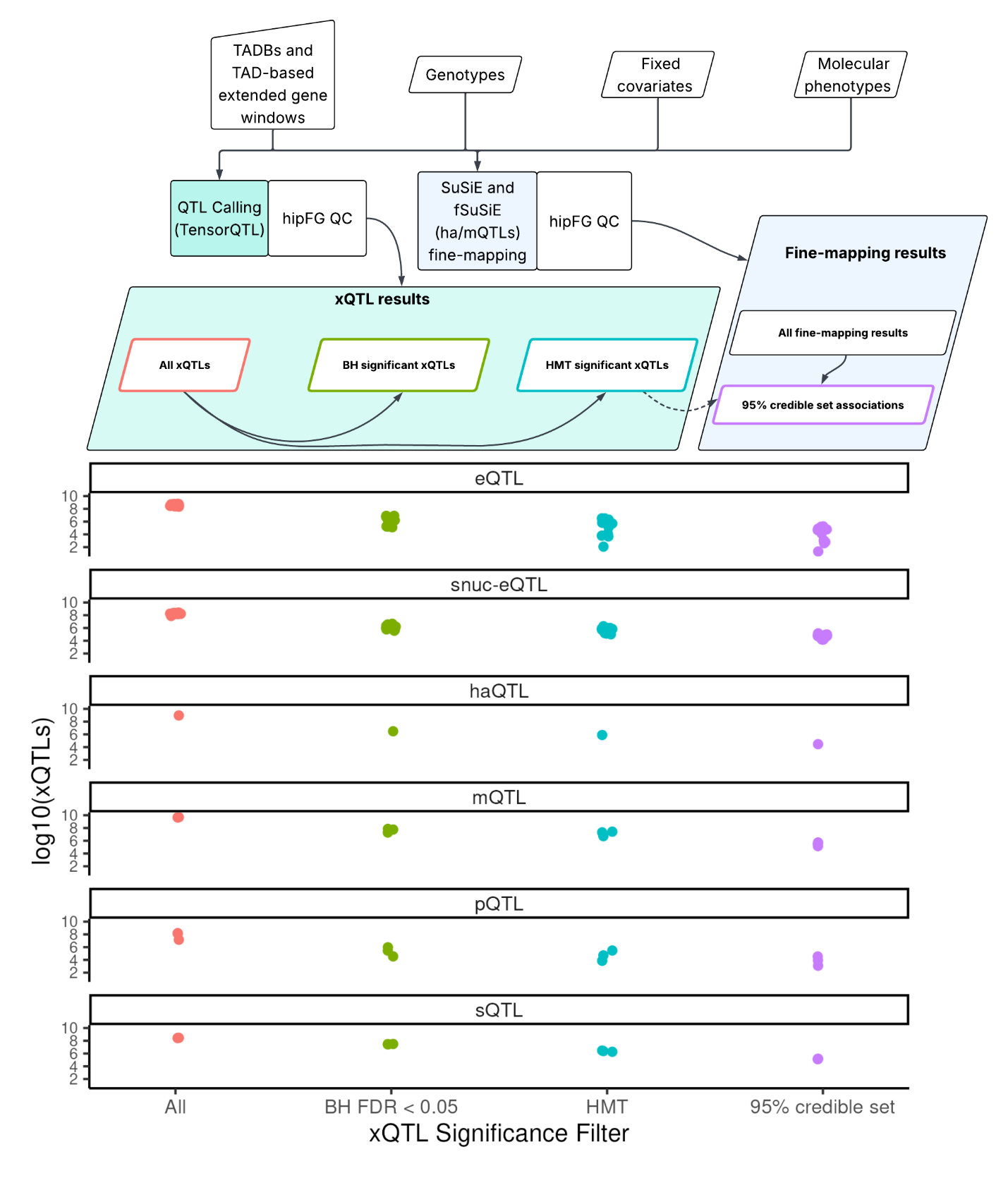

**Supplementary Fig. S3. Overview of released xQTL data products and file set composition. Top:** Overview of xQTL and fine-mapping association detection and significance-based filtering. All xQTLs: all xQTLs passing QC, with no significance requirements. BH-significant xQTLs: xQTLs with Benjamini-Hochberg (BH) multiple testing correction FDRs < 0.05 and nominal p-values < 0.01. HMT: xQTLs passing hierarchical multiple testing correction (HMT) (**Methods**). All fine-mapping results include SuSiE outputs without PIP or credible-set filters and fSuSiE outputs containing all associations in 95% credible sets. The 95% credible-set file includes associations in 95% credible sets after HMT-significant filtering. **Bottom:** Number of xQTLs in each dataset plotted with horizontal "jitter" for each significance level, separated by QTL type.

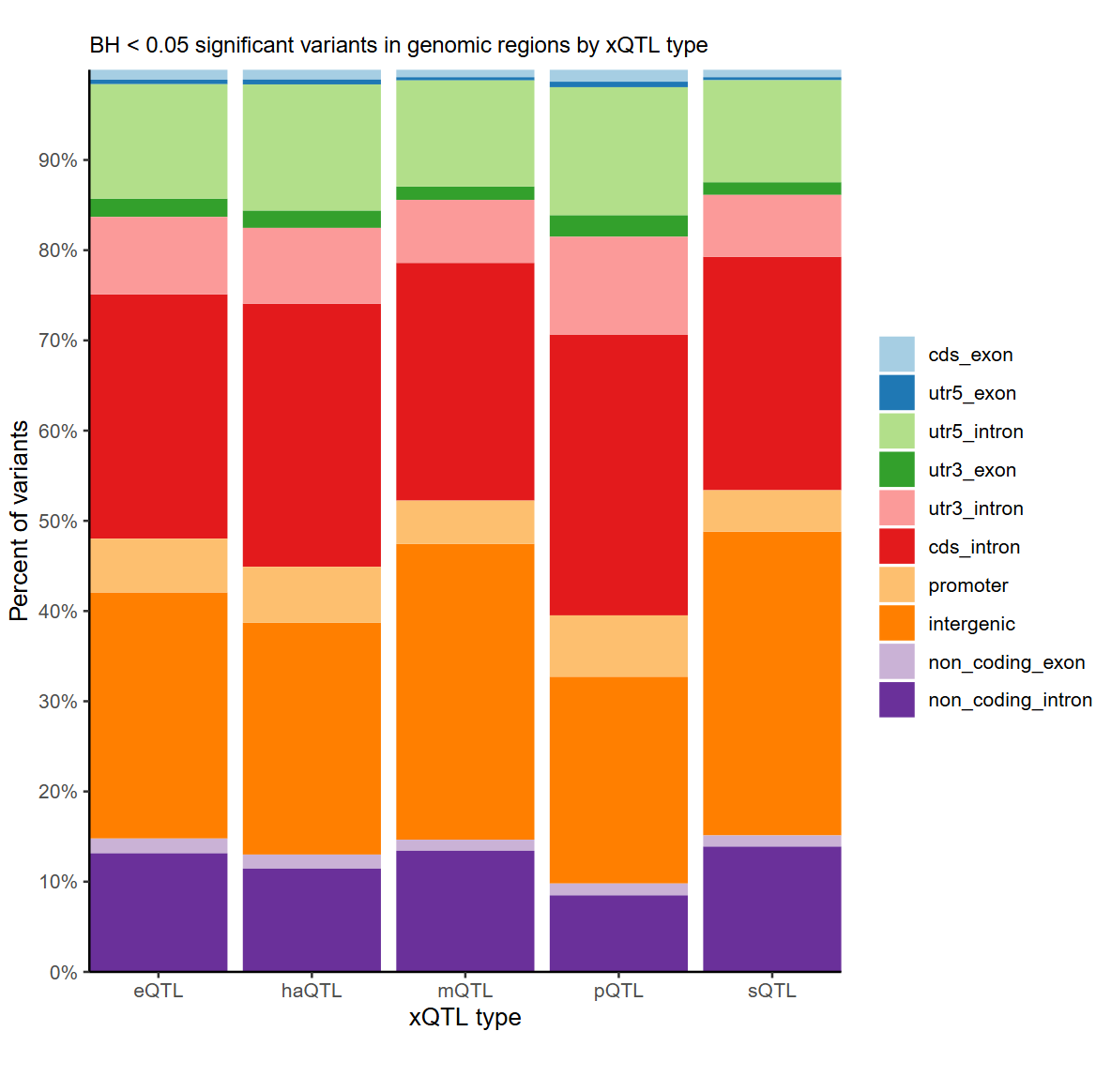
**Supplementary Fig. S4.** Genomic feature annotations of unique variants from BH-significant associations in each ROSMAP DLPFC xQTL dataset split by QTL type. Genomic feature categories are plotted and listed in the legend in descending priority order.

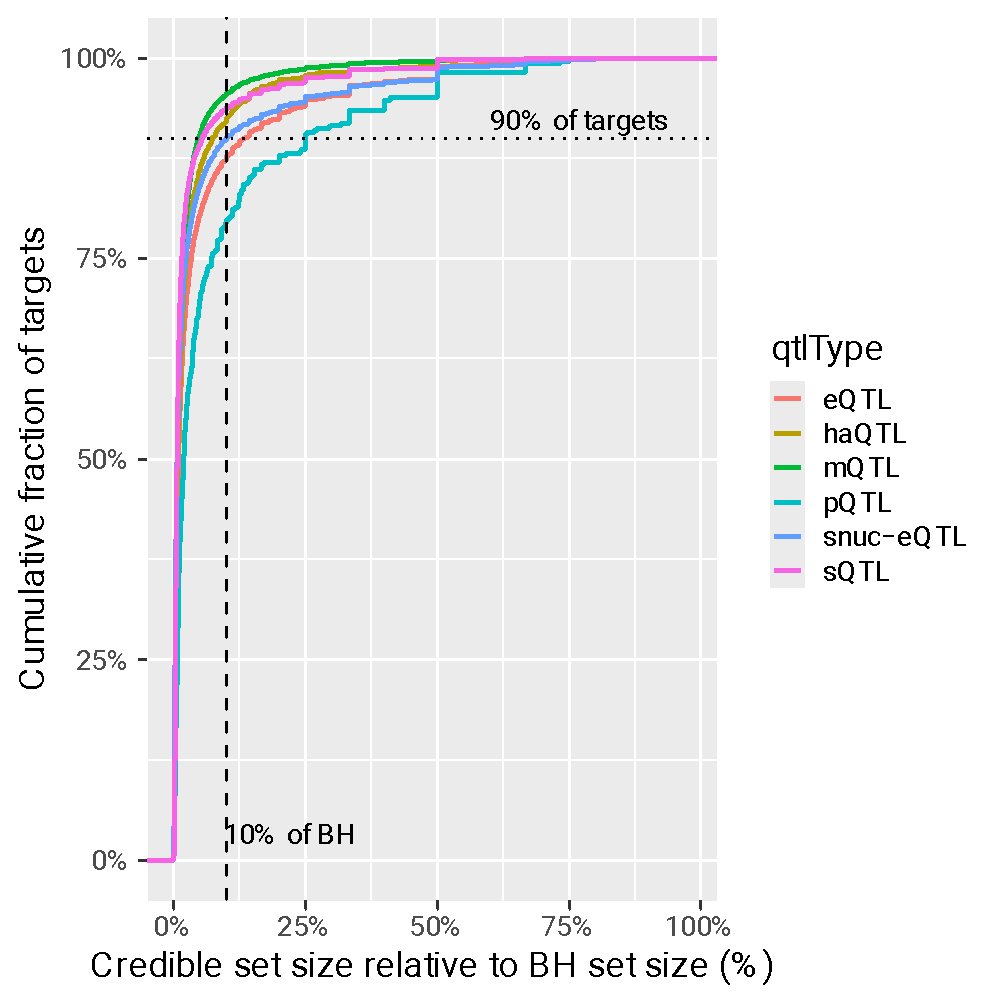

**Supplementary Fig. S5.** Credible set size reduction across QTL types. Empirical cumulative distribution function showing the size of fine-mapped credible sets relative to the corresponding Benjamini-Hochberg (BH) significant association set for each target. The x-axis represents credible set size as a percentage of the BH set size, and the y-axis represents the cumulative fraction of targets. Curves are stratified by QTL type, including eQTL, haQTL, mQTL, pQTL, snuc-eQTL, and sQTL. The dashed vertical line marks credible sets that are 10% of the BH set size, and the dashed horizontal line marks 90% of targets, highlighting the extent to which fine-mapping substantially reduces the number of candidate variants across molecular phenotypes.

### Supplementary References

1.	Cifello, J.*, et al.* hipFG: high-throughput harmonization and integration pipeline for functional genomics data. *Bioinformatics* **39**(2023).

2.	Yang, C.*, et al.* Genomic atlas of the proteome from brain, CSF and plasma prioritizes proteins implicated in neurological disorders. *Nat Neurosci* **24**, 1302-1312 (2021).

3.	Wang, M.*, et al.* The Mount Sinai cohort of large-scale genomic, transcriptomic and proteomic data in Alzheimer's disease. *Sci Data* **5**, 180185 (2018).

4.	Lopes, K.P.*, et al.* Genetic analysis of the human microglial transcriptome across brain regions, aging and disease pathologies. *Nat Genet* **54**, 4-17 (2022).

5.	De Jager, P.L.*, et al.* A multi-omic atlas of the human frontal cortex for aging and Alzheimer's disease research. *Sci Data* **5**, 180142 (2018).

6.	Genomes Project, C.*, et al.* A global reference for human genetic variation. *Nature* **526**, 68-74 (2015).

7.	Das, S.*, et al.* Next-generation genotype imputation service and methods. *Nat Genet* **48**, 1284-1287 (2016).

8.	Loh, P.R.*, et al.* Reference-based phasing using the Haplotype Reference Consortium panel. *Nat Genet* **48**, 1443-1448 (2016).

9.	Fernandez, M.V.*, et al.* Genetic and multi-omic resources for Alzheimer disease and related dementia from the Knight Alzheimer Disease Research Center. *Sci Data* **11**, 768 (2024).

10.	Li, H. A statistical framework for SNP calling, mutation discovery, association mapping and population genetical parameter estimation from sequencing data. *Bioinformatics* **27**, 2987-2993 (2011).

11.	Sherry, S.T.*, et al.* dbSNP: the NCBI database of genetic variation. *Nucleic Acids Res* **29**, 308-311 (2001).

12.	Purcell, S.*, et al.* PLINK: a tool set for whole-genome association and population-based linkage analyses. *Am J Hum Genet* **81**, 559-575 (2007).

13.	Manichaikul, A.*, et al.* Robust relationship inference in genome-wide association studies. *Bioinformatics* **26**, 2867-2873 (2010).

14.	Zhou, H.J., Li, L., Li, Y., Li, W. & Li, J.J. PCA outperforms popular hidden variable inference methods for molecular QTL mapping. *Genome Biol* **23**, 210 (2022).

15.	Zhou, W., Triche, T.J., Jr., Laird, P.W. & Shen, H. SeSAMe: reducing artifactual detection of DNA methylation by Infinium BeadChips in genomic deletions. *Nucleic Acids Res* **46**, e123 (2018).

16.	Chen, S. Ultrafast one-pass FASTQ data preprocessing, quality control, and deduplication using fastp. *Imeta* **2**, e107 (2023).

17.	Dobin, A.*, et al.* STAR: ultrafast universal RNA-seq aligner. *Bioinformatics* **29**, 15-21 (2013).

18.	Harrison, P.W.*, et al.* Ensembl 2024. *Nucleic Acids Res* **52**, D891-D899 (2024).

19.	Najar, C.*, et al.* Genetic and functional analysis of unproductive splicing using LeafCutter2. *bioRxiv* (2025).

20.	Hao, Y.*, et al.* Integrated analysis of multimodal single-cell data. *Cell* **184**, 3573-3587 e3529 (2021).

21.	Mathys, H.*, et al.* Single-cell multiregion dissection of Alzheimer's disease. *Nature* **632**, 858-868 (2024).

22.	Green, G.S.*, et al.* Cellular communities reveal trajectories of brain ageing and Alzheimer's disease. *Nature* **633**, 634-645 (2024).

23.	Ritchie, M.E.*, et al.* limma powers differential expression analyses for RNA-sequencing and microarray studies. *Nucleic Acids Res* **43**, e47 (2015).

24.	Gold, L.*, et al.* Aptamer-based multiplexed proteomic technology for biomarker discovery. *PLoS One* **5**, e15004 (2010).

25.	Seyfried, N.T.*, et al.* A Multi-network Approach Identifies Protein-Specific Co-expression in Asymptomatic and Symptomatic Alzheimer's Disease. *Cell Syst* **4**, 60-72 e64 (2017).

26.	Johnson, E.C.B.*, et al.* Large-scale proteomic analysis of Alzheimer's disease brain and cerebrospinal fluid reveals early changes in energy metabolism associated with microglia and astrocyte activation. *Nat Med* **26**, 769-780 (2020).

27.	Klein, H.U.*, et al.* Epigenome-wide study uncovers large-scale changes in histone acetylation driven by tau pathology in aging and Alzheimer's human brains. *Nat Neurosci* **22**, 37-46 (2019).

28.	Zhang, Y.*, et al.* Model-based analysis of ChIP-Seq (MACS). *Genome Biol* **9**, R137 (2008).

29.	Ernst, J. & Kellis, M. ChromHMM: automating chromatin-state discovery and characterization. *Nat Methods* **9**, 215-216 (2012).

30.	Lawrence, M.*, et al.* Software for computing and annotating genomic ranges. *PLoS Comput Biol* **9**, e1003118 (2013).

31.	Amemiya, H.M., Kundaje, A. & Boyle, A.P. The ENCODE Blacklist: Identification of Problematic Regions of the Genome. *Sci Rep* **9**, 9354 (2019).

32.	Ng, B.*, et al.* An xQTL map integrates the genetic architecture of the human brain's transcriptome and epigenome. *Nat Neurosci* **20**, 1418-1426 (2017).

33.	Consortium, G.T. The GTEx Consortium atlas of genetic regulatory effects across human tissues. *Science* **369**, 1318-1330 (2020).

34.	de Klein, N.*, et al.* Brain expression quantitative trait locus and network analyses reveal downstream effects and putative drivers for brain-related diseases. *Nat Genet* **55**, 377-388 (2023).

35.	Kerimov, N.*, et al.* A compendium of uniformly processed human gene expression and splicing quantitative trait loci. *Nat Genet* **53**, 1290-1299 (2021).

36.	Jia, Y.*, et al.* xQTLatlas: a comprehensive resource for human cellular-resolution multi-omics genetic regulatory landscape. *Nucleic Acids Res* **53**, D1270-D1277 (2025).

37.	Wang, G., Sarkar, A., Carbonetto, P. & Stephens, M. A simple new approach to variable selection in regression, with application to genetic fine mapping. *J R Stat Soc Series B Stat Methodol* **82**, 1273-1300 (2020).

38.	Denault, W.R.P.*, et al.* fSuSiE enables fine-mapping of QTLs from genome-scale molecular profiles. *bioRxiv* (2025).
